# Quantifying Localized Tumor-Infiltrating Lymphocytes by Deep Learning on H&E Images: Analytical Validation and Prognostic Value in Lung Adenocarcinoma

**DOI:** 10.64898/2026.10.23.26366340

**Authors:** Bolesław L. Osinski, Kshitij Ingale, Aileen Huang, Ben Terdich, Yoni Muller, Rohan Joshi, Riccardo Miotto, Ryan Jones, Nike Beaubier, Chithra Sangli

## Abstract

Tumor-infiltrating lymphocytes (TILs) are a critical determinant of response to immune checkpoint inhibitor (ICI) therapy in lung adenocarcinoma (LUAD). However, assessing the clinical utility of TIL quantification is challenging due to subjective manual scoring and differences in methodology, including some that may obscure localized immune responses critical to ICI response. Together, these issues limit our understanding of potential prognostic benefits of performing routine TIL quantification. Here, we present the “focal TIL score,” an artificial intelligence-powered biomarker derived from routinely generated Hematoxylin & Eosin (H&E) pathology images which measures TIL density within tiles across the entire relevant tissue region in order to account for spatial heterogeneity of localized lymphocytic infiltration. Underlying this method are two deep learning models that ensure accurate, reproducible, and interpretable quantification: one for lymphocyte detection trained on IHC-derived ground truth, and one for segmentation of tumor and tumor-adjacent stroma region (“scorable region”) trained on consensus pathologist annotations. We performed an analytical validation of the two models, demonstrating high overall performance: lymphocyte model concordance correlation coefficient (CCC) = 0.90, scorable region model Dice score = 0.87. We then performed a prognostic association study in a cohort of 685 ICI-treated LUAD patients and saw that, the focal TIL score was significantly associated with overall survival (HR=0.60, 95% CI 0.48-0.75, P<0.001), outperforming the standard biomarker PD-L1. Our findings show that quantifying the spatial intensity of immune infiltration, rather than the global average, provides a superior measure of the anti-tumor immune response. The accessibility and standardization associated with this approach may enable further studies and developments in precision oncology for LUAD.

## 1.0 Introduction

Lung cancer is the leading cause of cancer death in the US, with lung adenocarcinoma (LUAD) representing approximately 45% of all non-small-cell lung cancer (NSCLC) cases.^1,2^ For advanced-stage LUAD, immune checkpoint inhibitor (ICI) therapy, often in combination with chemotherapy, has become the standard treatment for most patients. Current biomarkers (such as PD-L1) enrich for response, but many patients do not respond to anti-PD-1 or anti-PD-L1 therapy.^3^ There is a critical need for scalable, tissue-based biomarkers to better capture the biology of the tumor immune microenvironment and more strongly predict patient response to ICI therapy—thereby ensuring that patients get put on the most effective therapy as quickly as possible. Routine pathology images provide a rich dataset, and leveraging these towards this goal need not alter clinician workflows.

The quantification of tumor-infiltrating lymphocytes (TILs) from hematoxylin & eosin (H&E) images has emerged as an alternative interpretable biomarker for predicting patient response to ICI therapy across many cancer types.^4–8^ Manual whole-slide TIL scoring methods, such as those standardized by the International TILs Working Group (TIL WG),^9^ have demonstrated prognostic value. However, manual estimation is inherently subjective and prone to inter-rater variability (Cohen’s kappa: 0.5–0.8), limiting its utility in clinical practice and significantly hampering studies into the overall efficacy of this biomarker.^10–12^

Machine learning approaches offer a robust, reproducible alternative to manual scoring. Previous work in triple-negative breast cancer demonstrated that an automated TIL quantification pipeline designed to follow the TIL WG guidelines could match the prognostic power of manual TIL WG scoring.^13^ Notably, this study used high quality IHC-derived ground truth labels for lymphocytes registered to H&E images. However, this study measured lymphocytes solely within tumor-associated stroma (TAS) regions. While this is a reasonable approach for resection specimens, in needle biopsies the tumor-stroma architecture is often not clearly defined. Consequently, the TIL WG and other more recent studies have recommended measuring TILs in combined tumor and stromal areas.^9,14–17^ One such study in NSCLC implemented a partially automated approach to measure TIL density within combined tumor and TAS regions, showing prognostic power and stratification within PD-L1 subgroups ^14^. While this study quantified TIL density through a single global averaging operation, other studies have demonstrated advantages to quantifying the spatial distribution of lymphocytes within the tumor microenvironment.^16,17^

Building on these advances, we developed and implemented the “focal TIL score”, a deep learning-based quantification method that is sensitive to localized regions of infiltration within tumor and TAS regions, known as immune “hotspots”^18,19^. Underlying the focal TIL score are two deep learning models: a lymphocyte detection model and a scorable region (tumor + TAS) segmentation model. We present a comprehensive analytical validation of both models including sub-analyses required for validation as a laboratory developed test. In a separate clinical cohort of ICI-treated LUAD patients reserved for prognostic association studies, we stratified patients as TIL High or Low (based on cutoffs for tile-level TIL density and slide-level infiltration fraction) and demonstrated that the focal TIL score is significantly associated with overall survival independent of clinical covariates. We also show that this association persists within PD-L1, tissue site, and surgical procedure strata. While the prognostic association analysis is framed as exploratory in absence of an independent external validation cohort, this preliminary evidence suggests that the H&E-derived focal TIL score could offer a standardized, interpretable biomarker for guiding LUAD treatment decisions.

## 2.0 Methods

### 2.1 Research ethics

All data for this study were pulled from the Tempus multimodal database (Tempus AI, Inc., Chicago, IL). This database contains de-identified, structured EHRs of oncology patients gathered from over 350 direct data connections across approximately 2,000 healthcare institutions in the United States that order Tempus products and services. This study and the data used received IRB exempt determinations (Advarra Pro00072742) as it was deemed to not be human subjects research; data were de-identified prior to analysis in accordance with the IRB exemptions.

### 2.2 I/E criteria for analytical validation cohort

We selected a total of 546 whole slide H&E images of LUAD patients from the de-identified Tempus multi-modal database for use in training either lymphocyte or scorable region models and analytical validation. Dataset curation was performed with the assistance of board-certified pathologists, who visually inspected each slide to ensure sufficient representation across all diagnostic subtypes of LUAD. Anatomic sites were restricted to lung/pleura, lymph node, liver, bone marrow, adrenal gland, and soft tissues. Within lung tissue, all adenocarcinoma diagnostic subtypes were represented including acinar, mucinous, solid, lepidic, papillary, and micropapillary. Samples deriving from fine needle aspirates and other discohesive cell preparations were excluded, as were slides derived from tissue that had undergone acid decalcification and tissues with less than 1 mm^2^ of tumor. Treatment-naive samples were preferentially selected. All whole slide images (WSIs) were scanned using a Leica Aperio GT 450 scanner. For inter-scanner variability analysis, all validation slides were additionally scanned on a Philips UFS scanner. For intra-scanner variability analysis, slides in the validation cohort were scanned on the same scanner type (Leica Aperio GT 450) on two different days; if the original section used for the analytical accuracy study was unavailable for rescan, an adjacent section was used.

### 2.3 I/E criteria for clinical exploration cohort

The clinical cohort used for prognostic association studies was drawn retrospectively from the de-identified Tempus Database and consisted of patients diagnosed with metastatic LUAD who received approved ICI as 1L therapy between January 2018, and July 2023. To be eligible, samples had to have an available H&E slide and sample collection had to occur before ICI exposure, within standard-of-care timing. Samples with discohesive tissue architectures (such as cytology specimens and fluid aspirates), tumor area < 1mm^2^, and Eastern Cooperative Oncology Group (ECOG) performance status score ≥3 were excluded. Metastases to bone were excluded, but all other tissue sites were allowed. For a full list of criteria see Supplementary Table S1. Patient characteristics of the clinical cohort matching these criteria (n=685) are shown in Table 1.

**Table 1.** Patient characteristics of the clinical cohort used for prognostic association studies.

|  | Count (percent) <sup>a</sup> |
| --- | --- |
| **Patient age** |  |
| < 65 | 292 (42.6%) |
| ≥ 65 | 393 (57.4%) |
| **Sex** |  |
| Female | 329 (48.0%) |
| Male | 356 (52.0%) |
| **Smoking status** |  |
| Non-smoker | 58 (8.5%) |
| Smoker | 602 (87.9%) |
| Unknown | 25 (3.6%) |
| **Stage at diagnosis** |  |
| Stage 1 | 20 (2.9%) |
| Stage 2 | 9 (1.3%) |
| Stage 3 | 28 (4.1%) |
| Stage 4 | 618 (90.2%) |
| Unknown | 10 (1.5%) |
| **Tumor area** |  |
| ≤ 5mm<sup>2</sup> | 357 (52.1%) |
| > 5mm<sup>2</sup> | 328 (47.9%) |
| **Procedure type** |  |
| Biopsy | 283 (41.3%) |
| Core needle biopsy | 284 (41.5%) |
| Excisional biopsy | 41 (6.0%) |
| Surgical resection | 77 (11.2%) |
| **Tissue site rollup** |  |
| Adrenal gland | 29 (4.2%) |
| Brain | 73 (10.7%) |
| Liver | 56 (8.2%) |
| Lung | 299 (43.6%) |
| Lymph node | 97 (14.2%) |
| Pleura (solid tissue) | 47 (6.9%) |
| Other <sup>b</sup> | 84 (12.3%) |
| **Regimen** |  |
| Atezolizumab | 15 (2.2%) |
| Nivolumab | 44 (6.4%) |
| Pembrolizumab | 609 (88.9%) |
| Other | 17 (2.5%) |
| **Therapy regimen type** |  |
| Unknown | 1 (0.1%) |
| Combo | 584 (85.3%) |
| Mono | 100 (14.6%) |
| **Scanner make** |  |
| Aperio | 386 (56.4%) |
| Philips | 299 (43.6%) |
| **TMB status** |  |
| High | 234 (34.2%) |
| Low | 451 (65.8%) |
| **PD-L1 status** |  |
| Negative | 225 (32.8%) |
| Positive | 401 (58.5%) |
| Unknown | 59 (8.6%) |

<sup>a</sup> Percentages are out of total clinical cohort size (n=685)
<sup>b</sup> Over 20 distinct tissue sites with low prevalence are rolled into “Other” (see SI)

### 2.4 PD-L1 and TMB status

PD-L1 status was determined by IHC testing at Tempus or curated pathology reports from external PD-L1 IHC testing at referring labs. PD-L1 positive and negative classifications were defined according to standardized FDA guidelines for NSCLC, namely tumor proportion score (TPS) ≥ 1%, applicable across various PD-L1 clones.

Genomic profiling was performed using the Tempus xT DNA sequencing assay.^20,21^ TMB was calculated as the number of nonsynonymous variations divided by the respective panel size (detailed in Supplementary Text S.1).

### 2.5 Ground truth for lymphocyte model

Out of the 546 samples selected for model development, 280 samples with unstained sections were reserved for lymphocyte model development. Previous studies have shown considerable variability in manual annotation of lymphocytes from H&E images, and recommend using IHC as a more objective ground truth.^10,13,22^ Because consecutive H&E and IHC tissue sections will not contain the same cells, we opted to stain H&E and IHC on the same section. To do so, unstained sections were initially selected in order to minimize the time between H&E staining and de-staining, as the de-staining process works best on fresh stains. These were stained for H&E and scanned, then immediately shipped to an external vendor (San Diego Pathology) to be de-stained with xylene and re-stained with CD3/CD20 IHC (T-Cell/B-Cell lymphocytes), after which they were sent back and scanned again. This produced a paired dataset of H&E and IHC stained images on the same exact tissue (Figure 1A).

**Figure 1.**
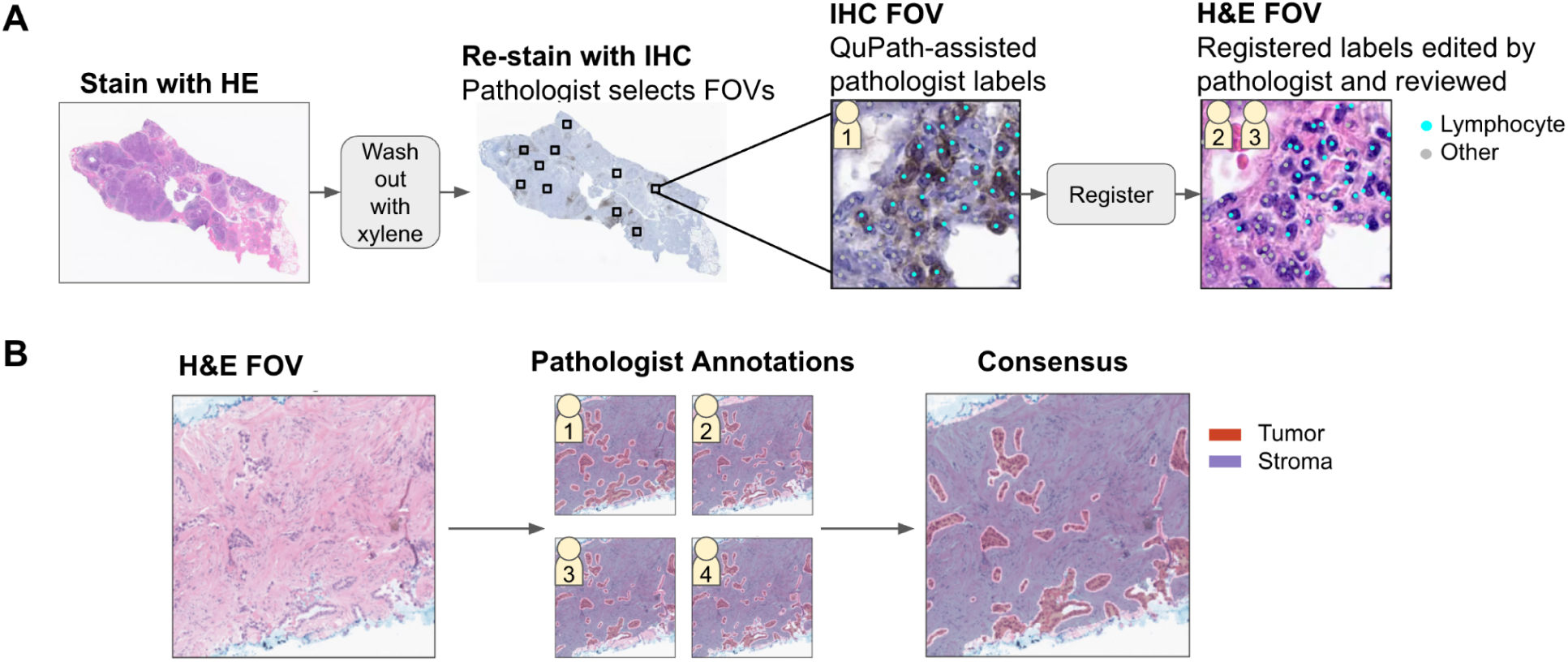
**(A)** Ground truth collection workflows for lymphocytes. One pathologist selects FOVs on the IHC image and labels stained lymphocytes in QuPath. Cell labels are registered to the corresponding H&E image, where a second pathologist fixes labeling and registration errors. Due to the difficulty of the task, a 3rd pathologist makes final corrections of the cell labels. **(B)** Ground truth annotations for tumor and stroma regions are performed on each FOV by 4 pathologists. Consensus is then computed by a voting scheme.

Using publicly available software, QuPath,^23^ pathologists annotated 5-10 fields of view (FOVs) on each IHC slide and ran stain detection to label each cell as Lymphocyte or Other. The QuPath cell detection coordinates were then registered to the corresponding H&E using a custom multi-resolution iterative closest points algorithm. Slides with poor IHC staining or completely failed registration were removed, reducing the dataset to 228 slides. These data were split into train (110) / tuning (56) / test (62) sets. For slides in the training set, FOVs were cropped to 128×128μm^2^ (containing a maximum of ∼700 cells), while for slides in the tuning/test sets FOVs were cropped to 64×64μm^2^ (containing a maximum of ∼90 cells). Larger FOVs for training ensured higher numbers at the cost of label accuracy, while the smaller FOVs for tuning and testing ensured that each cell could be visually reviewed by a pathologist to maximize accuracy (Supplementary Figure S1).

Even for the slides where registration was successful, we observed subtle tissue warping which accumulates to a considerable shift at the scale of single cells (Supplementary Figure S2). Therefore, despite performing IHC and H&E staining on the same exact tissue, perfect automated registration of cell labels was impossible for most slides. Errors could also arise from the QuPath cell detection on IHC, which struggled to label cells especially in areas of densely packed lymphocytes. To address these issues a 2nd pathologist edited the registered labels on the H&E image, followed by a 3rd review from another pathologist to make additional corrections as needed. Pathologist assignments were randomized such that no pathologist reviewed their own edits and each FOV had input from 3 pathologists. For model training and evaluation, FOVs were read from WSIs at 20x magnification.

### 2.6 Ground truth for tumor and stroma region model

Out of the 546 samples selected for model development, 266 H&E stained WSIs were selected for region model development. Tumor and stroma region annotations were performed directly on H&E slides within FOVs (1mm^2^) by 4 pathologists per FOV, after which the consensus was computed by a voting scheme (Figure 1B). The data were split into train (140) / tuning (60) / test (66) sets. In addition, previously collected annotations on 60 LUAD slides were used for intermediate evaluation of the trained model. For model training and evaluation, FOVs were read from WSIs at 20x magnification.

For the test set we asked the pathologists not to annotate tumor and stroma regions, but instead to annotate the “scorable region”, defined as the tumor region and 50μm of tumor-associated stroma extending beyond the tumor border. This was chosen so that we evaluate performance on the final endpoint of the region prediction pipeline.

### 2.7 Definition of scorable area

We defined the “scorable area” as the tumor region combined with a 50 μm margin of surrounding tumor-associated stroma (TAS). This margin width was selected to capture lymphocytes actively infiltrating the tumor border—a feature known to correlate with outcomes^8,9^—while excluding distant tertiary lymphoid structures. Literature regarding optimal margin width varies widely (ranging from 32 μm to 500 μm).^9,12^ Some authors suggest simply measuring TILs on the entire tissue,^15^ and our own internal sensitivity analysis found that TIL density is in fact largely insensitive to margin variations between 50 μm and 250 μm (Supplementary Figure S3). Therefore, the exact choice of margin width is not likely to have a large impact on results.

To generate the scorable area, region model predictions were post-processed as follows: (1) tumor masks were filtered to remove regions smaller than 0.0032 mm² (approximately 5 tumor cells) to eliminate spurious predictions; (2) the cleaned tumor mask was dilated by 50 μm; (3) this dilated region was intersected with the predicted stroma mask to isolate the TAS region, while excluding distant stroma; (4) the union of cleaned tumor and TAS regions is taken.

### 2.8 Focal TIL scoring pipeline

We developed a fully-automated focal TIL scoring pipeline consisting of five steps, depicted in Figure 2A. Briefly, the tissue is detected on an H&E image and the scorable region and lymphocyte models are run in parallel on the tissue. The predicted scorable region is then tiled into 1 mm^2^ tiles. For each tile, the density of lymphocytes is computed within the enclosed scorable region and a cutoff for TIL density is applied to identify infiltrated tiles. Next, the slide-level fraction of infiltrated tiles is measured, which is reported as the focal TIL score. A slide-level cutoff then assigns the sample a TIL High or TIL Low status. Examples of the pipeline predictions for needle biopsy and surgical resection specimens are shown in Figure 2B.

**Figure 2.**
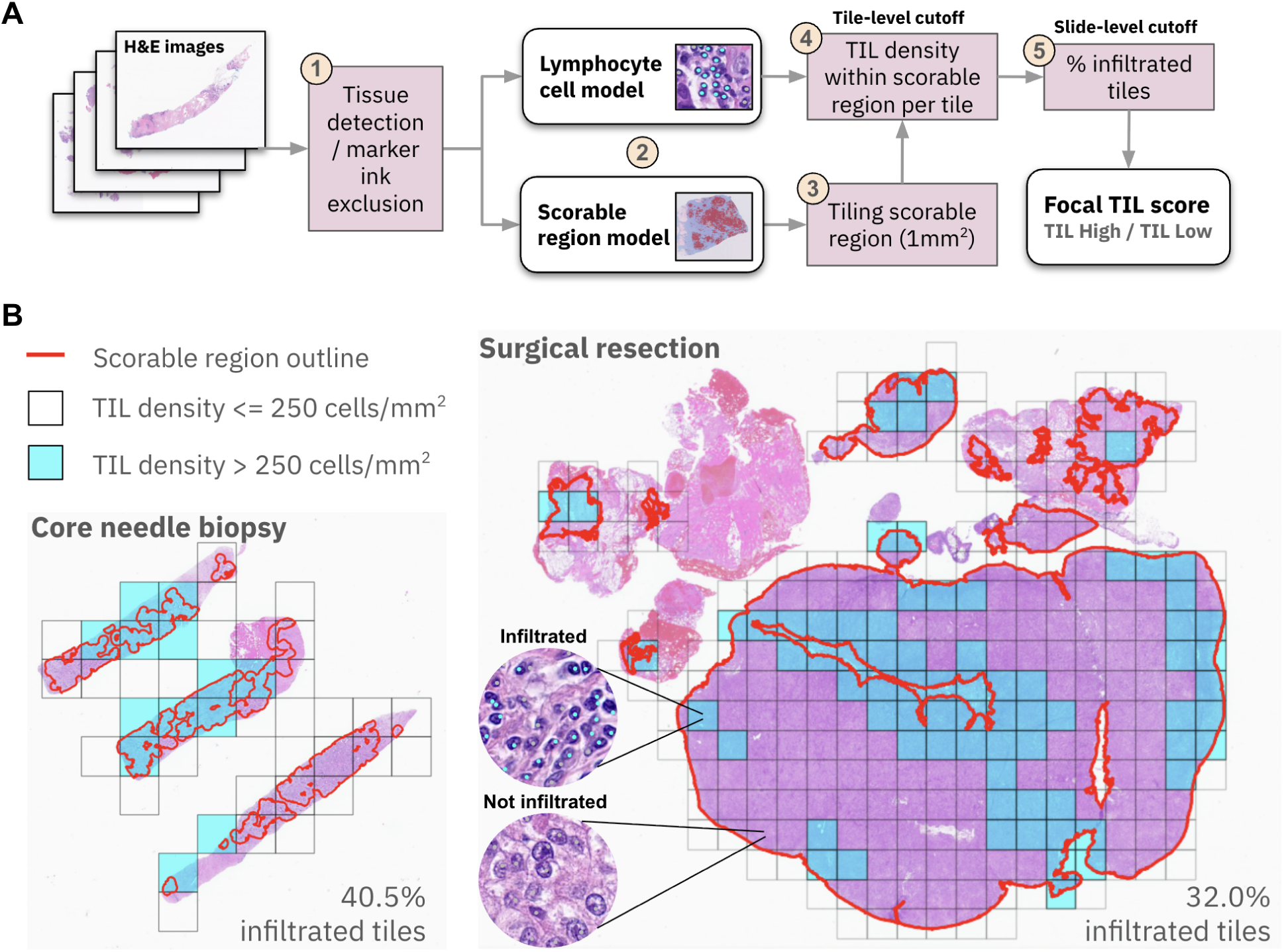
**(A)** Fully-automated focal TIL scoring pipeline design. (1) Two previously developed models for detecting tissue and marker ink at 0.625x magnification restrict analysis to suitable tissue regions. (2) A U-net segments tumor and stroma regions from within the tissue, which is then post-processed to create the scorable region (tumor + TAS). In parallel, a second cell-level U-net detects lymphocytes. (3) The scorable region is tiled into 1 mm^2^ tiles. (4) For each tile, the lymphocyte density is measured in the scorable region within the tile, and a cutoff is applied to determine if the tile is infiltrated. (5) The focal TIL score is reported as a percentage of infiltrated tiles. A second slide-level cutoff is applied to this percentage to assign a TIL High or TIL Low category. **(B)** Examples of focal TIL measurements on core needle biopsy (left) and surgical resection (right) specimens. The scorable region is outlined in red. Tiles with TIL densities above the tile-level TIL density cutoff are colored cyan. Representative infiltrated and non-infiltrated regions are shown with cell model predictions overlaid. Localization of immune hotspots is especially visible in the resection specimen. Slide-level infiltration fractions are shown as percentages.

As opposed to many of the published TIL quantification methods in the literature, which use a single cutoff to assign TIL scores,^6,7,12–15^ this two-cutoff approach provides an additional degree of freedom to calibrate sensitivity to localized regions of immune infiltration (hotspots).

### 2.9 Cell detection model

**Model Architecture:** A 5-layer residual U-net architecture^24^ was trained via strong supervision to detect lymphocytes using point annotations. To adapt the segmentation architecture for point-based supervision, we utilized a masked cross-entropy loss function. Point annotations were converted into pseudo-masks consisting of a central circular region (calibrated to the 7-10 um diameter of a lymphocyte) surrounded by an ignored annular region, which together created a “donut” shaped mask (Supplementary Figure S4). This design penalizes loss only within the cell center while disregarding uncertain boundary regions. Although optimized for lymphocyte dimensions, this masking strategy proved effective for the “other cell” class as well.

**Training and Optimization:** Training was performed with data augmentation including color jitter, H&E stain normalization, and spatial transformations. Standard hyperparameters were tuned to maximize the lymphocyte F1 score on the hold-out tuning set (final parameters: Adam optimizer, learning weight 1e-3, and weight decay 1e-3). Class weighting and the pseudo-mask generation kernel size were additionally tuned to maximize Concordance Correlation Coefficient (CCC), balance precision and recall, and resolve closely packed cells. The optimal values for these parameters were: lymphocyte class weight = 100; other cell class weight = 50; mask kernel size = 5×5 (Supplementary Figure S5).

### 2.10 Tumor and stroma region model

**Model Architecture:** A 6-layer U-net with 3 residual units was trained via strong supervision with standard cross-entropy loss to segment tumor and stromal compartments.

**Training and Optimization:** Weights were initialized by the used Kaiming He method.^25^ To enhance model robustness, the primary LUAD training set was supplemented with annotations from a metastatic breast cancer cohort (N=308). This co-training strategy significantly improved performance compared to training on LUAD alone, likely due to shared morphological features across tissue types (Supplementary Table S2). Hyperparameters were tuned to maximize the combined Dice score for tumor and stroma classes. The final model utilized the Adam optimizer with a learning rate of 1e-6 and weight decay of 1e-4.

### 2.11 Statistical methods for analytical validation

Cell detection performance was evaluated using the classification F1 score and the CCC of cell counts. The F1 score was calculated from a global confusion matrix accumulated across all FOVs (detailed in Supplementary Text). Continuous predicted probabilities from U-net were converted to discrete points located at peak probabilities (Supplementary Figure S6, Supplementary Table S3). These predicted cells were matched to ground truth centroids using a fixed radius of 6μm (detailed in Supplementary Text). To compute CCC, the number of ground truth cells and predicted cells within each FOV are summed respectively and then correlated. While F1 measures individual detection accuracy, CCC was employed to quantify systematic bias (under- or over-estimation) by comparing predicted and ground truth cell counts per FOV against the line of identity (x=y).

Region segmentation performance was evaluated using the Dice similarity coefficient, a standard measure of spatial overlap that is mathematically equivalent to the F1 score.^26^

### 2.12 Statistical methods for prognostic association

Cutoffs for tile-level TIL density and slide-level infiltration fraction were determined via a 100-fold cross-validated hazard ratio minimization procedure (detailed in Supplementary Text S.4, Supplementary Figure S7). Patients were classified as TIL High if tile-level TIL density was > 250 cells/mm^2^ and slide-level infiltration fraction was > 0.30.

Prognostic association was performed to measure the association of focal TIL score (high vs. low) with overall survival (OS), defined as the time from ICI start date to death from any cause. Event-free patients were censored at the date of last known follow-up or at the end of the study period of 24 months, whichever occurred first. Although all sample biopsies were performed prior to ICI treatment, subsequent data collection (imaging and sequencing) could have occurred before or after ICI initiation. To account for delayed entry, where imaging/sequencing occurred after ICI initiation, all analyses incorporated a risk-set adjustment where the entry time was defined as the later of imaging/sequencing date or ICI start date.

Prognostic association analyses were performed using the Kaplan-Meier estimator and the Cox Proportional Hazards model (lifelines v0.30.0; Python 3.10). Hazard ratios (HR) and 95% confidence intervals (CI) were reported, with significance evaluated using a one-sided 5% Wald test. Univariate and multivariate analyses were performed to assess the association between focal TIL score and OS independent of clinical factors (age < 65, sex, smoking status, tumor size, surgical procedure type, treatment regimen type) and known biomarkers (TMB and PD-L1 status). Subgroup analyses were stratified by tissue site and PD-L1 status. Multivariate analysis was restricted to samples with complete clinical data, reducing the population from 685 to 626. No multiplicity adjustments were performed.

## 3.0 Results

### 3.1 Analytical accuracy and robustness

The analytical performance of the lymphocyte and scorable region models (on their respective withheld test sets) are presented in Table 2. Across the full test set, the lymphocyte model achieved CCC=0.90 and F1=0.74, while the region model achieved Dice=0.87. Performance varied by tissue site: lymphocyte detection was most accurate in lung (F1=0.78) and lymph node (F1=0.76) samples, followed by moderate performance in soft tissue, liver, and adrenal gland (F1=0.48∼0.53). Performance was notably lower in bone tissue (F1=0.23), likely due to the presence of confounding hematopoietic elements that mimic the appearance of lymphocytes. For this reason, bone tissue was excluded from the clinical dataset used for subsequent prognostic association. The scorable region model, on the other hand, performed robustly across all tissue sites (Dice=0.87∼0.91). Both models performed well across all procedure types and diagnostic subtypes of LUAD.

**Table 2.** Summary of Analytical Validation of Lymphocyte and scorable region models.

| Characteristic | Cohort Sizes |  | Analytical Validation Results |  |  |
| --- | --- | --- | --- | --- | --- |
|  | Lymphocyte<br>N slides (N<br>FOVs) | Scorable<br>region<br>N slides (N<br>FOVs) | Lymphocyte<br>CCC <sup>a</sup> | Lymphocyte<br>F1 <sup>b</sup> | Scorable<br>Region Dice <sup>c</sup> |
| **Tissue Site** |  |  |  |  |  |
| Lung | 33 (313) | 32 (63) | 0.93 | 0.78 | 0.87 |
| Lymph node | 13 (119) | 13 (24) | 0.92 | 0.76 | 0.87 |
| Liver | 3 (25) | 5 (10) | 0.38 | 0.50 | 0.91 |
| Bone | 4 (35) | 5 (10) | 0.17 | 0.23 | 0.92 |
| Adrenal gland | 5 (47) | 6 (12) | 0.59 | 0.53 | 0.85 |
| Soft tissue | 4 (40) | 5 (9) | 0.81 | 0.48 | 0.88 |
| **Procedure Type** |  |  |  |  |  |
| Resection &<br>Excision | 15 (150) | 11 (22) | 0.95 | 0.82 | 0.91 |
| Core needle<br>biopsy | 47 (429) | 55 (107) | 0.87 | 0.70 | 0.86 |
| **Subtype (lung**<br>**tissue)** |  |  |  |  |  |
| Acinar | 10 (88) | 10 (20) | 0.87 | 0.67 | 0.88 |
|  | Lymphocyte<br>N slides (N<br>FOVs) | Scorable<br>region<br>N slides (N<br>FOVs) | Lymphocyte<br>CCC <sup>a</sup> | Lymphocyte<br>F1 <sup>b</sup> | Scorable<br>Region Dice <sup>c</sup> |
| **Tissue Site** |  |  |  |  |  |
| Lung | 33 (313) | 32 (63) | 0.93 | 0.78 | 0.87 |
| Lymph node | 13 (119) | 13 (24) | 0.92 | 0.76 | 0.87 |
| Mucinous | 9 (90) | 8 (16) | 0.94 | 0.76 | 0.86 |
| Solid | 6 (60) | 5 (10) | 0.83 | 0.67 | 0.84 |
| Lepidic | 2 (15) | 3 (6) | 0.92 | 0.86 | 0.74 |
| Papillary | 2 (20) | 0 (0) | 0.97 | 0.91 | N/A |
| Micropapillary | 2 (20) | 2 (4) | 0.97 | 0.81 | 0.96 |
| Unspecified | 2 (20) | 4 (7) | 0.93 | 0.71 | 0.90 |
| **Total (all tissues)** | **62 (579)** | **66 (128)** | **0.90** | **0.74** | **0.87** |

<sup>a</sup> CCC: concordance correlation coefficient. Measures deviation from x=y unity line. Useful for measuring bias in under/over counting, but not appropriate for low cell count regimes (such as liver).
<sup>b</sup> F1 lymphocyte classification: Measures cell-to-cell classification performance computed in bulk from a single confusion matrix, which is summed from all FOVs in the given characteristic. This is a more appropriate metric for tissues with small numbers of lymphocytes.
<sup>c</sup> Dice score (F1) is computed from a single bulk confusion matrix which is summed from all FOVs in the given characteristic.

The low CCC values in bone and adrenal gland tissue stemmed from a positive bias (over-counting) relative to ground truth (Supplementary Figure S8). In contrast, the low CCC observed in liver tissue likely reflects a limitation of the metric in sparse data regimes rather than poor model performance. Given the physiological scarcity of lymphocytes in the liver,^28^ the low variance in cell counts renders CCC unstable. Detailed inspection revealed that cell count predictions were accurately low in 96% (24/25) of FOVs, but the poor CCC was driven by a single outlier (Supplementary Figure S8). Thus, the model accurately reflected the “immune desert” phenotype typical of liver tissue.

We also demonstrated robustness of both models to scanner differences and artifacts. Model predictions were highly concordant when evaluated on Leica Aperio GT450 and Philips UFS scans of the same H&E slides (lymphocyte CCC=0.99, scorable region CCC=1.0, Supplementary Figure S9A) as well as between two scans of the same slides scanned months apart on Leica Aperio GT450 scanners (lymphocyte CCC=0.99, scorable region CCC=1.0, Supplementary Figure S9A). To quantify robustness to artifacts, we performed a simulation where a random percentage of image tiles were artificially perturbed with either random color augmentation or Gaussian blurring. We measured the maximum allowable artifact percentage as the maximum percentage of tiles satisfying the following acceptance criteria between images with and without the perturbations: lymphocyte cell count CCC>0.95, scorable region F1>0.85. For the lymphocyte model, the maximum allowable percentage for color perturbation and blurring was 20% (Supplementary Figure S9B). For the scorable region model, the maximum allowable percentage was 20% for color perturbation, and 50% for blurring (Supplementary Figure S9C).

### 3.2 Univariate and multivariate analysis of focal TIL association with overall survival

To assess the prognostic value of the focal TIL score, we performed Cox Proportional Hazards modeling (with risk set adjustment) on a cohort of patients receiving first-line ICI treatment (Figure 3A, Supplementary Table S4). Patients were stratified using the cutoffs defined in Methods (tile-level > 250 cells/mm^2^; slide-level > 0.30). In univariate analyses, the focal TIL score demonstrated the strongest association with OS (HR=0.60, CI 0.47-0.76, p<0.001). Other variables including PD-L1 positivity (HR=0.68, CI 0.54-0.86, p<0.01) and age < 65 years (HR=0.69, CI 0.54-0.88, p<0.01) were also significantly associated with longer survival in univariate analyses. Conversely, needle biopsy procedure type was associated with shorter survival compared to surgical resection (HR=1.50; 95% CI 1.1–2.1; p=0.017), a finding likely attributable to the preferential use of biopsies in advanced, unresectable disease.

**Figure 3.**
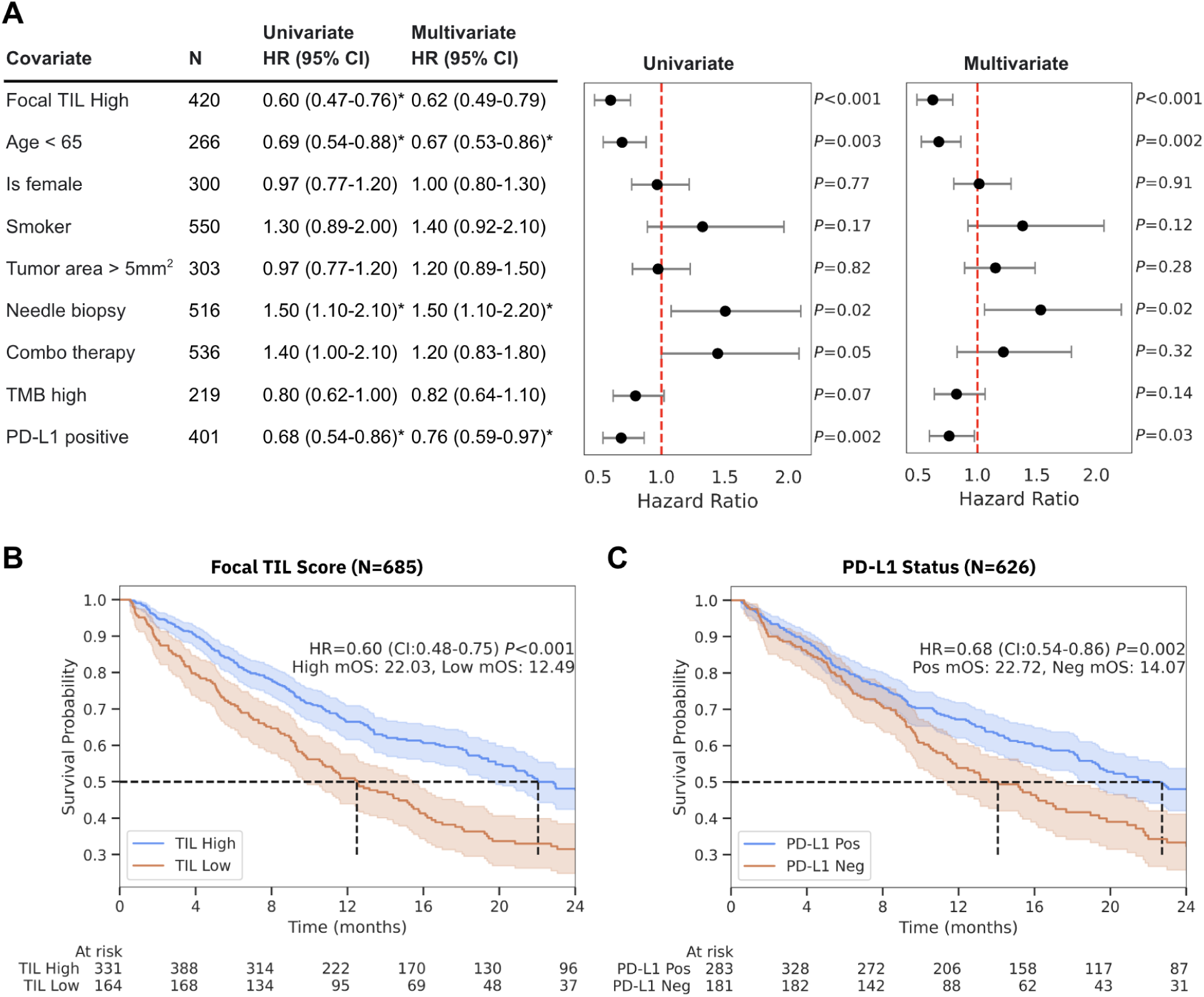
Association of Focal TIL score and clinical covariates with overall survival (OS). **(A)** Univariate (one fit per covariate) and multivariate (fit on all covariates) Cox Proportional Hazards analysis of the focal TIL score alongside standard clinical factors and biomarkers. The dataset was restricted to samples with completed data (N=626). The table (left) and corresponding forest plots (right) display Hazard Ratios (HR) and 95% Confidence Intervals (CI). Factors with statistically significant associations (p < 0.05) are marked with asterisks. In the forest plots, the red dashed line represents the null hypothesis (HR=1.0). **(B)** Kaplan-Meier survival curves stratified by Focal TIL score (High vs. Low) in the entire clinical dataset (N=685). Shaded areas represent 95% confidence intervals. Hazard ratios, p-values, and median overall survival (mOS) in months are annotated for each comparison. Dashed black lines indicate the median survival time. Note that due to risk set adjustment accounting for delayed entry, the “At risk” counts do not monotonically decrease. **(C)** Kaplan-Meier survival curves stratified by PD-L1 status (Positive vs. Negative) in the restricted dataset (N=626). Same figure layout as (B).

To evaluate the independence of the focal TIL score, a risk set adjusted multivariate Cox regression was performed including all factors in a single fit (Supplementary Table S5). The focal TIL score remained a significant independent predictor of OS (HR=0.62, CI 0.49-0.79, p<0.001). Similarly, age < 65 (HR=0.67, CI 0.53-0.86, p=0.002), PD-L1 positivity (HR=0.76, CI 0.59-0.97, p=0.03), and needle biopsy procedure type (HR=1.5, CI 1.1-2.2, p=0.02) maintained their significance. TMB status, sex, smoking status, tumor area, and treatment regimen were not significantly associated with outcome in either univariate or multivariate models.

Univariate analysis was also performed to assess associations between tissue site and OS (Supplementary Table S6). Metastasis to the “Other” tissue category (n=75, which contains > 20 mixed sites that are listed in Supplementary Table S7) was associated with shorter survival (HR=1.4, CI 1.0-2.0, p=0.03). Unexpectedly, metastasis to the brain (n=67) was associated with longer survival (HR=0.42, CI 0.25-0.7, p<0.001). No significant associations were found for the remaining tissue sites.

Kaplan-Meier survival curves further visualized the risk stratification of the focal TIL score in the full clinical dataset (N=685; Figure 3B). Patients classified as TIL High had a significantly longer median overall survival (mOS) of 22.0 months compared to 12.5 months for the TIL Low group (HR=0.60, CI 0.48-0.75, p<0.001). By comparison, stratification based on PD-L1 status (Figure 3C, restricted to samples with PD-L1 status, n=626) yielded a mOS of 22.7 months for PD-L1 positive versus 14.1 months for PD-L1 negative patients (HR=0.68, CI 0.54-0.86, p=0.002).

### 3.3 Subgroup Analysis: PD-L1 Status, Tissue Site, and Procedure Type

To evaluate the prognostic value of the focal TIL score independent of PD-L1 status, we performed Kaplan-Meier analysis and fit univariate Cox PH models within PD-L1 positive and negative subgroups (Figure 4A). The focal TIL score maintained significant risk stratification in both the PD-L1 positive (HR=0.61, CI 0.45-0.84, p=0.002) and PD-L1 negative (HR=0.65, CI 0.45-0.93, p=0.019) cohorts, indicating that the focal TIL score provides prognostic information distinct from and additive to PD-L1 expression.

**Figure 4.**
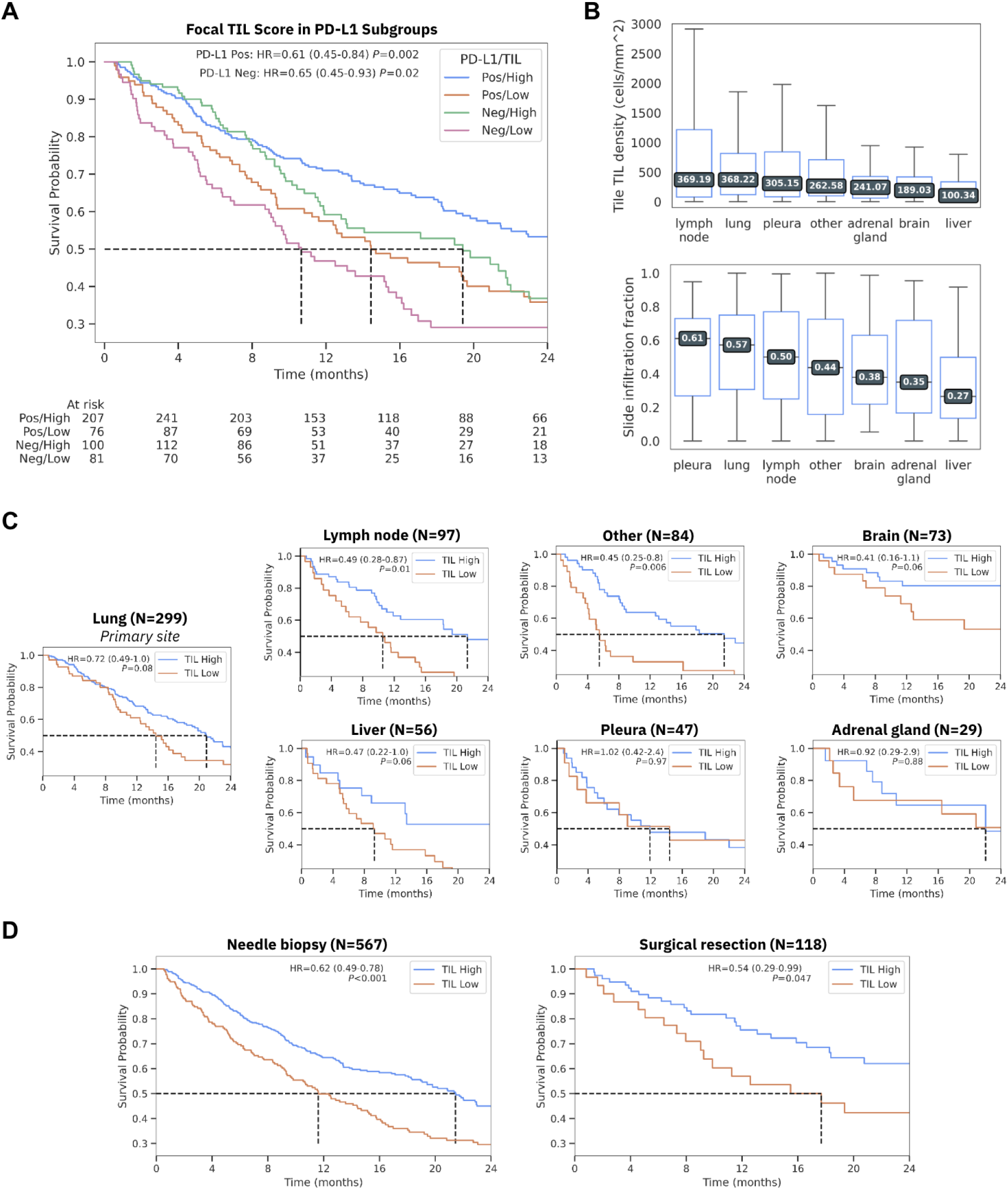
Prognostic robustness of Focal TIL score across clinical and anatomical subgroups. **(A)** In risk set adjusted Kaplan-Meier analysis, the focal TIL score significantly stratifies risk within both PD-L1 Positive (HR=0.61, p=0.002) and PD-L1 Negative (HR=0.65, p=0.019) cohorts. **(B)** Distribution of immune infiltration metrics across tissue sites. Top: Tile-level TIL density (cells/mm2). Bottom: Slide-level infiltration fraction. Boxes represent the interquartile range; values inside boxes indicate the median. Lymph node and lung tissues exhibit the highest median TIL density, while liver exhibits the lowest. **(C)** Kaplan-Meier curves evaluating the focal TIL score within tissue site subgroups consisting of the primary site (lung), and six metastatic sites. Hazard ratios (HR) and p-values are annotated. **(D)** Kaplan-Meier curves within procedure type subgroups, demonstrating significant prognostic value of the focal TIL score in both needle biopsies (n=567, p<0.001) and surgical resections (n=118, p=0.047).

We further characterized the immune landscape across distinct anatomical sites (Figure 4B). As expected, lymph node and lung samples exhibited the highest median tile-level TIL densities (369 and 368 cells/mm^2^, respectively), while liver metastasis samples showed the lowest infiltration (100 cells/mm^2^). Despite this heterogeneity in baseline density, the focal TIL score showed association with OS across several tissue sites (Figure 4C). Significant stratification was observed in lymph node metastases (HR=0.49, p=0.014, n=97) and the “Other” tissue category (HR=0.45, p=0.006, n=84). Notably, the prognostic power in primary lung tissue, while directionally consistent (HR=0.72), did not reach statistical significance (p=0.08, n=299). Similarly, directionally consistent but non-significant signals persisted in liver (HR=0.47, p=0.06, n=56) and brain (HR=0.41, p=0.06, n=73) sites. No association with OS was observed in pleura (n=47) or adrenal gland (n=29) subgroups, though this may be driven by their smaller sample sizes.

Finally, consistent with the need for a biomarker suitable for diverse clinical workflows, we assessed prognostic performance by procedure type (Figure 4D). The focal TIL score significantly stratified OS in the needle biopsy subgroup (HR=0.62, p<0.001, n=567), which represents the majority of advanced-stage diagnoses. Importantly, significant stratification was also achieved in the surgical resection subgroup (HR=0.54, p=0.047, n=118). This robustness to tissue size distinguished the focal methodology from standard approaches; in a direct comparison, a global averaging TIL score failed to stratify risk within the resection subgroup (p>0.05), likely due to signal dilution in larger tissue areas (Supplementary Figure S10). Thus, unlike global averaging, the focal TIL score maintains prognostic utility regardless of the total tissue surface area.

## 4.0 Discussion

### Summary & Mechanism of Action

In this study, we presented the “focal TIL score”, a spatially resolved method for quantifying TILs in H&E images. We performed a rigorous analytical validation of the lymphocyte detection and scorable region segmentation models underlying the method, showing strong performance of both models across all diagnostic subtypes of LUAD along with robustness to artifacts and scanner differences. The performance of both of these models is in line with other state-of-the-art models.^12,27^ We also demonstrated that the focal TIL score significantly associates with OS in patients with advanced LUAD treated with ICI.

The stronger prognostic association with OS of the focal TIL score compared to TMB and PD-L1 likely reflects its role as a direct measure of the effector immune response. While TMB serves as an upstream proxy for potential immunogenicity, high mutational load does not guarantee T-cell infiltration due to potential defects in antigen presentation or trafficking.^29^ Similarly, while PD-L1 expression is the current clinical standard, it remains an imperfect predictive biomarker due to inconsistencies in assay platforms and scoring protocols, and an inability to distinguish between “hot” and “cold” immune infiltration states.^30^ By quantifying high-density immune hotspots within tumor regions, the focal TIL score acts as a downstream measure of the immune cycle, identifying an infiltration phenotype that is primed to respond to checkpoint blockade.

### Methodological Advantage: Focal vs. Global

We acknowledge that the prognostic value of TILs in NSCLC has not always outperformed PD-L1 in prior literature.^31^ We postulate that this discrepancy may stem from the global averaging method in standard TIL scoring protocols. Global averaging can dilute the signal of localized immune responses, particularly in large tissue sections where heterogeneous “cold” regions obscure active hotspots. Crucially, in a direct comparison within our cohort, we observed that global averaging significantly underestimated TIL infiltration in larger tumors compared to the focal score, and failed to stratify outcomes in surgical resections (Supplementary Figure S10). Thus our findings suggest that the added spatial context of the focal TIL score improves clinical utility for specimens with larger tissue surface areas.

### Clinical Use Cases

The focal TIL score could offer utility across multiple distinct clinical scenarios in the management of cancer treatment. The score could serve as a predictive biomarker to identify patients most likely to respond to ICIs, such as pembrolizumab or nivolumab, thereby refining patient selection beyond PD-L1 status alone. It could also help tailor therapeutic intensity: immune infiltrated tumors might be candidates for toxicity-sparing ICI monotherapy, whereas immune deserts may require combinatorial regimens (e.g., chemotherapy-IO) to overcome resistance. Additionally, for emerging Adoptive Cell Therapies (ACT) such as TIL therapy (e.g., lifileucel), this TIL quantification method could act as a crucial screening tool to assess eligibility or guide the selection of T-cell-rich tissue blocks for therapeutic harvesting.

### Complementarity to NGS for immune deconvolution

Our findings also highlight the value of imaging-based biomarkers in contexts where next-generation sequencing (NGS) faces limitations. While NGS is the gold standard for TMB and neoantigen load, bulk sequencing methods struggle to resolve tumor-specific immune signals in lymph node metastases, where the high background of native lymphocytes confounds immune deconvolution. ^32^ Imaging methods inherently preserve spatial context, allowing for the segmentation of tumor regions from background lymphoid tissue. The robust analytical performance and prognostic signal we observed in lymph node metastases suggests that the focal TIL score may serve as a critical complement to genomic profiling in determining the immune status of nodal disease, offering more robust multimodal ICI-prediction.

### Performance in Metastatic Sites & Subgroups

An unexpected finding was that prognostic stratification was often more robust in metastatic sites than in primary lung tissue. This is notable given that our analytical validation indicated lower cell detection performance in complex metastatic tissues (e.g., liver, bone, soft tissue) compared to lung. It is possible that the primary lung samples in the clinical cohort had complex morphologies (e.g. lepidic) that interfered with TIL quantification, but this was not visually assessed. However, this discrepancy implies that the biological signal of immune infiltration in metastatic sites is sufficiently potent to drive risk stratification despite higher analytical noise. It suggests that perfect cell-level precision is not a prerequisite for capturing the macro-level clinical signal (High vs. Low), supporting the utility of the focal TIL score across diverse metastatic contexts.

### Limitations

Our study has limitations. Firstly, while we performed a rigorous cross-validation procedure to derive robust cutoffs to binarize the focal TIL score, these cutoffs require validation on external, independent cohorts to confirm their generalizability. Notably, our selected cutoffs do align with existing literature (250 cells/mm² and 0.30 infiltration fraction).^13,16^ But in the absence of a withheld clinical test set, our prognostic analysis can only be considered exploratory and not a proper clinical validation. Secondly, Our current model does not explicitly quantify tertiary lymphoid structures (TLS), which are known to coordinate potent anti-tumor responses.^18,33^ Thirdly, our model was only trained and analytically validated in LUAD, leaving squamous cell carcinomas out of the assessment. Finally, we also noted a counter-intuitive finding that brain metastases were associated with longer OS. This could reflect selection bias inherent to retrospective clinical cohorts. Patients with brain metastases are typically excluded from clinical trials or immunotherapy treatment regimens unless their CNS disease is stable or has been treated with stereotactic radiosurgery.^34^ Thus, this subgroup likely represents a selected population with controlled intracranial disease, rather than reflecting the full population of LUAD metastasis to the brain.

### Future Directions

While we achieved acceptable analytical performance using standard strongly supervised deep learning architectures, generalizing this strongly supervised modeling approach across a wide range of cancer indications and tissue sites (e.g. bone) would be challenging and inefficient due to the tedious and costly nature of ground truth collection for lymphocytes. The emergence of computational pathology foundation models trained on massive, pan-cancer datasets offers a promising pathway to address this.^35,36^ By leveraging these pre-trained encoders, future iterations of TIL quantification tools may achieve the robust generalization across all tissue histologies required for broad clinical adoption. Besides generalizing the TIL model across indications, future iterations of the model could aim to explicitly capture other signatures of immune infiltration, such as TLS. In addition, advances in high-resolution spatial transcriptomics offer spatially resolved labels of cell subtypes on H&E images, which could be used to train future TIL models to identify specific lymphocyte subtypes, such as cytotoxic T cells. While this study was retrospective, a future prospective clinical trial should be run using focal TILs to drive therapy decisions (with or without PD-L1) in order to evaluate its true effectiveness.

## Data Availability

All data produced in the present study are available upon reasonable request to the authors

## Acknowledgements

We thank Alia Zander, Rossin Erbe, and Ailin Jin for assistance with risk set adjusted prognostic analyses. We also thank Adam Hockenberry and Dana DeSantis for their editorial assistance in preparing the manuscript.

