## Supplementary Information for "Quantifying Localized Tumor-Infiltrating Lymphocytes by Deep Learning on H&E Images: Analytical Validation and Prognostic Value in Lung Adenocarcinoma"

### Supplementary Tables

**Table S1. Clinical cohort I/E criteria**

| Inclusion Criteria | Exclusion criteria |
|---|---|
| • Received ICI treatment as 1L in the advanced setting<br>• Metastatic lung adenocarcinoma diagnosis<br>• Metastatic diagnosis date is < 07/01/2023<br>• At least 18 years old at metastatic diagnosis<br>• At least 1 day of follow-up after treatment start<br>• De novo stage IV or metastatic recurrence at time of sample collection<br>• Time between sample collection date and LOT1 initiation must be <=90 days<br>• Time between metastatic diagnosis and LOT1 initiation must be <=90 days<br>• Time between the start of LOT and the start of ICI must be <=60 days<br>• Scorable area > 1mm | • Overlap with cell or region model development sets<br>• Patients that received an investigational new drug as part of their ICI regimen<br>• Patients with more than 1 year between initiation of ICI and sample collection<br>• Patients with a known ECOG PS ≥ 3 where ECOG PS was assessed after metastatic diagnosis and within 30 days after treatment start<br>• Tissue based pathologic diagnosis not confirmed within 60 days of clinically reported metastatic diagnosis by imaging<br>• Patients with pathogenic mutations in EGFR<br>• Patients with fusions involving ALK, ROS1, RET, or NTRK1/2/3<br>• Cytology specimen or discohesive tissue architecture (as assessed by pathologist)<br>• Bone tissue sites<br>• Out of focus H&E slide image |

**Table S2. Region model improves with co-training on NSCLC and Breast**

| | Test set F1 (DICE) scores for Tumor (N=60) | | | | Test set F1 (DICE) scores for Stroma (N=60) | | | |
|---|---|---|---|---|---|---|---|---|
| | All | Lung | Lymph Node | Other mets* | All | Lung | Lymph Node | Other mets |
| Train on HE NSCLC (N=140 slides) | 0.78 | 0.70 | 0.86 | 0.85 | 0.66 | 0.65 | 0.68 | 0.64 |
| Co-train on NSCLC & breast (N=448 slides) | 0.85 | 0.79 | 0.90 | 0.89 | 0.78 | 0.78 | 0.75 | 0.81 |

* Other mets includes liver, bone, soft tissue, and adrenal gland

**Table S3. Summary of distance parameters used to train and evaluate the lymphocyte detection model**

| Parameter name | Used for | Values (pixels at 20x mag) | Effective size in um |
|---|---|---|---|
| Donut masks for cross entropy | Converting point annotations to pseudo-masks for U-net training | Kernel 5x5,<br>Inner iteration = 1<br>Outer iteration = 2 | Inner radius = 6um<br>Outer radius = 12um |
| Min distance for peaks (radius) | Convereting predicted continuous probabilities back to points | R = 5 | 2*R+1 = 6um |
| Assignment distance (radius) | Assigning predicted points to ground truth annotated points | R = 6 | 2*R+1 = 7um |

**Table S4. Univariate analysis table**

| Cobariate | HR (95% CI) | p | N within group | N outside group | mOS* within group (months) | mOS outside group (months) |
|---|---|---|---|---|---|---|
| Focal TIL High | 0.60 (0.47-0.76) | 0.000018 | 420 | 206 | 22.85 | 12.85 |
| Age < 65 | 0.69 (0.54-0.88) | 0.002575 | 266 | 360 | 23.93 | 15.97 |
| Is female | 0.97 (0.77-1.2) | 0.767413 | 300 | 326 | 18.95 | 19.41 |
| Smoker | 1.30 (0.89-2.0) | 0.16687 | 550 | 76 | 18.33 | >24.0 |
| Tumor area > 5mm$^2$ | 0.97 (0.77-1.2) | 0.817182 | 303 | 323 | 18.26 | 19.44 |
| Needle biopsy** | 1.50 (1.1-2.1) | 0.017057 | 516 | 110 | 18.13 | >24.0 |
| Combo therapy | 1.40 (1.0-2.1) | 0.050903 | 536 | 90 | 18.26 | >24.0 |
| TMB high | 0.80 (0.62-1.0) | 0.072411 | 219 | 407 | 22.72 | 16.62 |
| PD-L1 positive | 0.68 (0.54-0.86) | 0.001527 | 401 | 225 | 22.72 | 14.07 |

**Table S5. Multivariate analysis table**

| Covariate | HR (95% CI) | p | N within group | N outside group |
|---|---|---|---|---|
| Focal TIL High | 0.62 (0.49-0.79) | 0.000118 | 420 | 206 |
| Age < 65 | 0.67 (0.53-0.86) | 0.001528 | 266 | 360 |
| Is female | 1.00 (0.80-1.30) | 0.906469 | 300 | 326 |
| Smoker | 1.40 (0.92-2.10) | 0.117871 | 550 | 76 |
| Tumor area > $5mm^2$ | 1.20 (0.89-1.50) | 0.279109 | 303 | 323 |
| Needle biopsy** | 1.50 (1.10-2.20) | 0.023018 | 516 | 110 |
| Combo therapy | 1.20 (0.83-1.8) | 0.315177 | 536 | 90 |
| TMB high | 0.82 (0.64-1.10) | 0.139124 | 219 | 407 |
| PD-L1 positive | 0.76 (0.59-0.970) | 0.030513 | 401 | 225 |

* mOS >24 indicates that the survival probability did not fall below 0.5 before the maximum time was reached (24 months).
** Some samples were labeled only as “biopsy” in clinical notes. These were grouped into needle biopsy after visual inspection.

**Table S6. Univariate analysis for tissue sites**

| Tissue site | HR (95% CI) | p | N within group | N outside group | mOS within group (months) | mOS outside group (months) |
|---|---|---|---|---|---|---|
| **Lung** | 0.86 (0.68-1.10) | 0.197297 | 274 | 352 | 19.80 | 18.26 |
| **Lymph node** | 1.20 (0.85-1.60) | 0.331968 | 86 | 540 | 15.48 | 19.44 |
| **Brain** | 0.42 (0.25-0.70) | 0.000749 | 67 | 559 | >24.0 | 18.07 |
| **Liver** | 1.50 (1.00-2.10) | 0.050212 | 52 | 574 | 11.61 | 19.80 |
| **Pleura** | 1.40 (0.95-2.20) | 0.087724 | 45 | 581 | 10.72 | 19.41 |
| **Adrenal gland** | 0.79 (0.43-1.40) | 0.437079 | 27 | 599 | >24.0 | 19.05 |
| **Other** | 1.40 (1.00-2.00) | 0.030309 | 75 | 551 | 13.08 | 19.44 |

Metastasis to some tissue sites may be predictive of OS. Tissue sites with significant association with OS are brain, liver, and other metastatic sites. Curiously, metastasis to brain is significantly associated with longer survival, while metastasis to the other tissue category is significantly associated with shorter survival. Metastasis to liver is directionally associated with shorter survival, though not significantly. Due to relatively small sample sizes in these groups (brain N=67, other N=75, liver N=52), these effects may not hold in a larger population.

**Table S7 Composition of the "Other" Tissue SIte Category:** In the prognostic analysis cohort, the "Other" category (N=75) comprises over 20 distinct metastatic sites with low individual sample counts. The exact list of all tissue sites included in this category is shown in the table.

| Tissue site | Count |
|---|---|
| Thorax or connective, subcutaneous and other soft tissues of thorax | 24 |
| Head, face or neck | 6 |
| Other ill-defined sites | 5 |
| Connective, subcutaneous and other soft tissues of lower limb and hip | 5 |
| Lower limb | 4 |
| Connective, subcutaneous and other soft tissues of trunk | 4 |
| Mediastinum | 4 |
| Abdomen | 4 |
| Upper limb | 3 |
| Breast | 3 |
| Skin of scalp and neck | 3 |
| Connective, subcutaneous and other soft tissues of abdomen | 2 |
| Unknown | 2 |
| Connective, subcutaneous and other soft tissues of upper limb and shoulder | 2 |
| Skin of trunk | 2 |
| Specified parts of peritoneum | 1 |
| Heart | 1 |
| Kidney | 1 |
| Pancreas | 1 |
| Jejunum | 1 |
| Connective, subcutaneous and other soft tissues of head, face, and neck | 1 |
| Testis | 1 |
| Ill-defined sites within respiratory system | 1 |
| Skin of other and unspecified parts of face | 1 |
| Connective, subcutaneous and other soft tissues of pelvis | 1 |
| Esophagus | 1 |

### Supplementary Figures

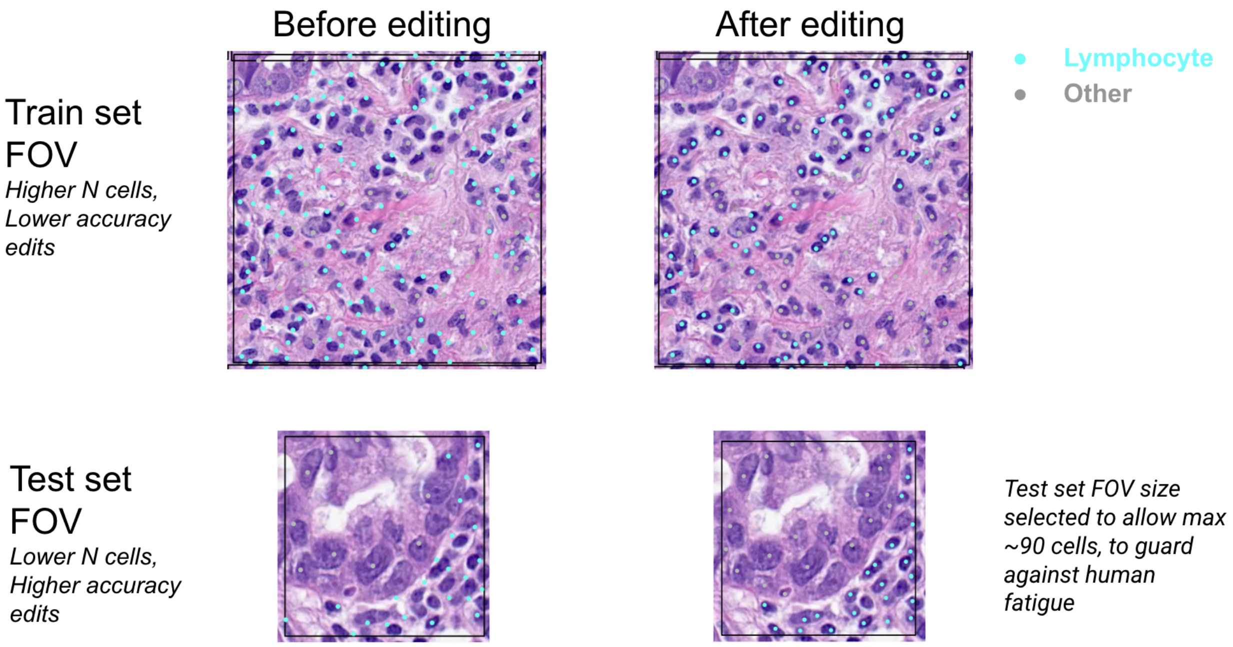


**Figure S1. IHC-derived cell labels registered to H&E must still be manually edited due to subtle registration errors**.

While registration from IHC to H&E sometimes results in good alignment of cell labels, we found that for the majority of slides the labels were slightly shifted. The figure shows and example of QuPath-derived cell labels registered from IHC to HE before (left) and (after) right

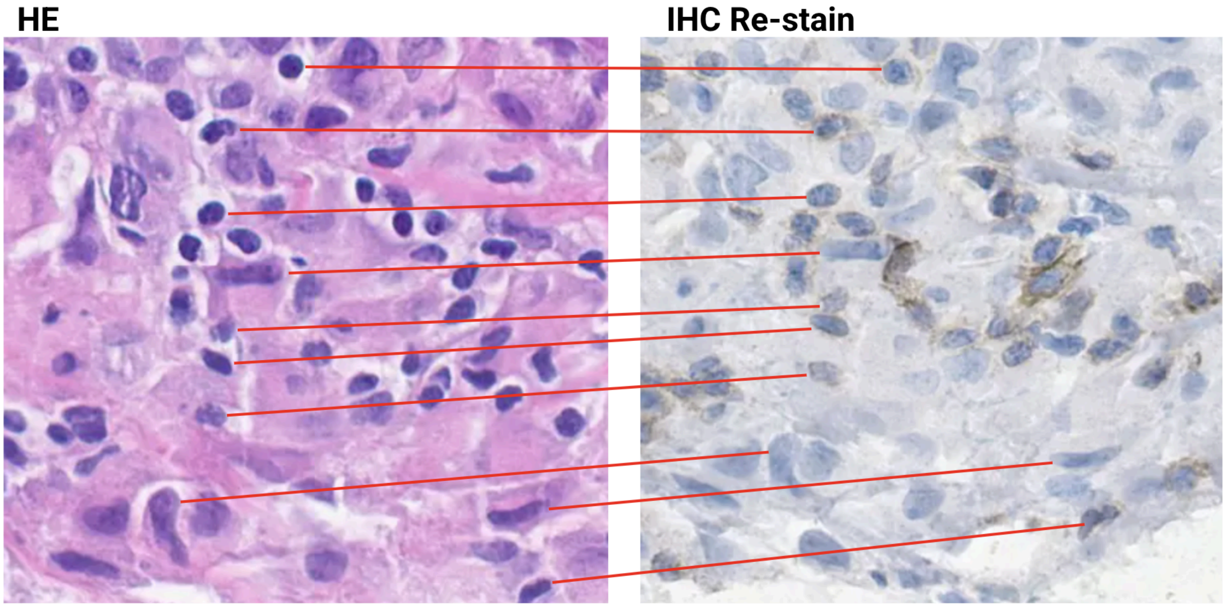


**Figure S2. Cumulative warp of IHC re-stained tissue relative to initial H&E stain.**
Even though the IHC is re-stained on the exact same tissue as the H&E, we found that the IHC staining procedure can cause distortions such as shrinkage and displacements which warp the IHC relative to the H&E. The figure shows corresponding regions (at the same magnification) in H&E and IHC stained tissues. Cells are aligned at the top, but due to the cumulative warp the cells towards the bottom of the IHC image are shifted upwards relative to the H&E image.

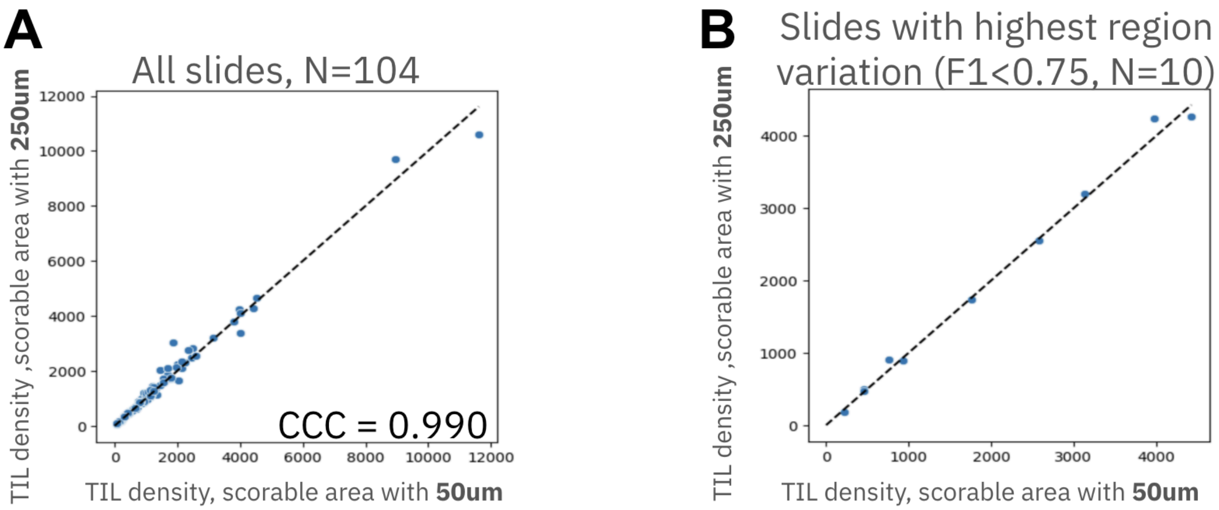


**Figure S3. TIL density is largely insensitive to changes in margin width**
**(A)** In an exploratory dataset (non-overlapping with analytical and clinical cohorts), the standard global averaged TIL density (not using tiling) was computed from the scorable regions defined with either 50µm or 250µm TAS margin widths. A scatter plot shows that TIL density is insensitive to margin width. **(B)** This robustness holds even when restricted to slides with highest region variation between the scorable areas defined with the two margin widths (when region F1<0.75). This stability can be due to a combination of the following factors: 1) In biopsy samples the margin is often not present, so irrelevant, 3) the tumor area is much larger than the margin so the contribution of margin is small, 3) the density of lymphocytes is relatively constant through the margin.

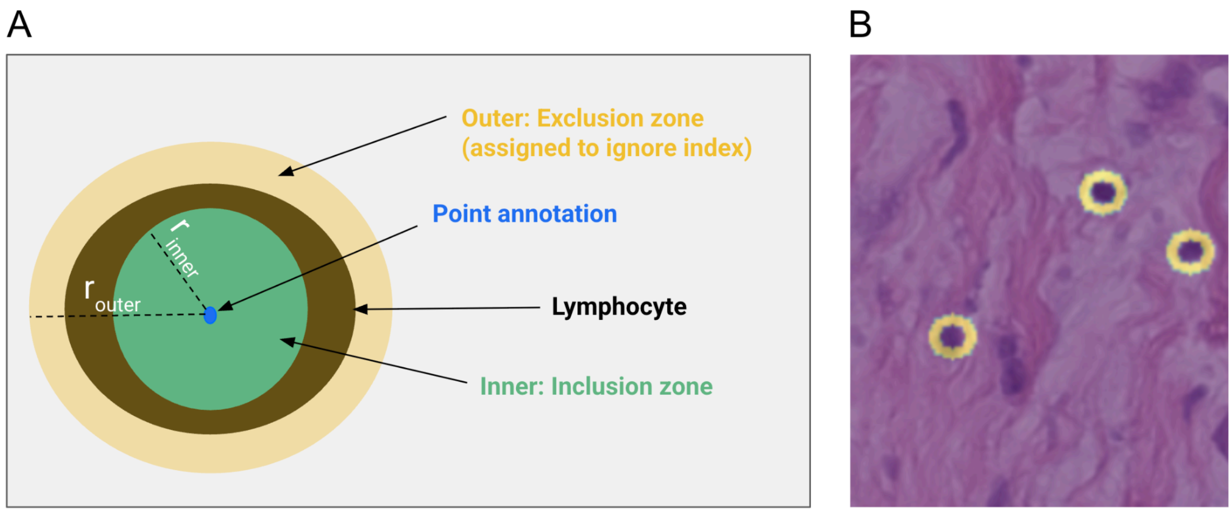


**Figure S4. Method for creating point annotation pseudo-masks that enable training of UNet model. (A)** Schematic depicting point masking by concentric regions. This creates a "donut" shape, characterized by an inner radius, $r_{inner}$, which is slightly smaller than an average lymphocyte nucleus and an outer radius, $r_{outer}$, extending beyond the edge on an average lymphocyte nucleus. The outer annular region defines an exclusion zone where cross entropy is ignored for training a UNet model. The mask is created by dilating the point masks using cv2.MORPH_ELIPSE with a 5x5 kernel, 1 iteration for the inner zone, and 2 iterations for the exclusion zone. **(B)** Pseudo-masks overlaid on three lymphocytes at 20x magnification, demonstrating masks matching the scale of a lymphocyte, typically 7-10um in diameter.

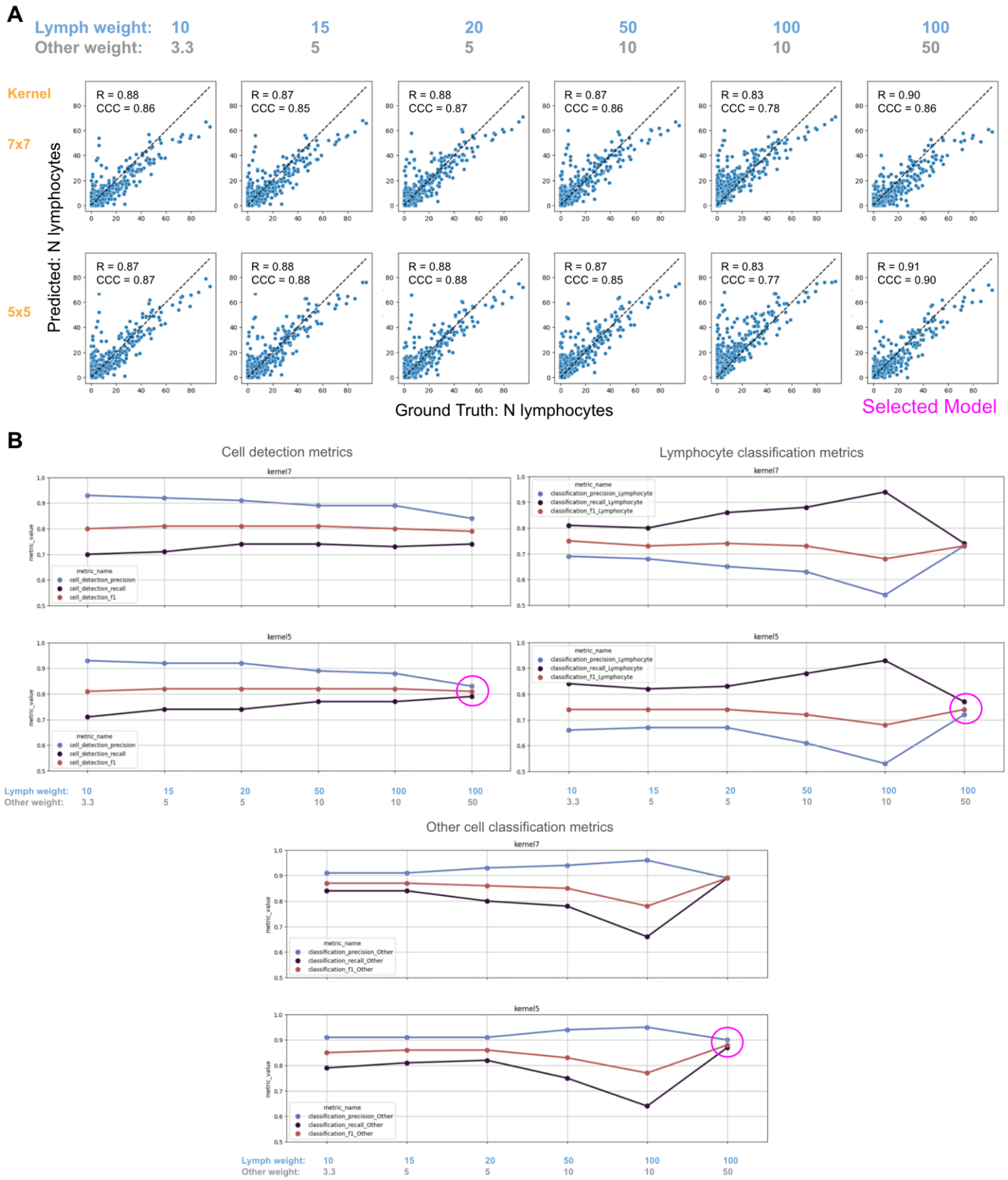


**Figure S5. Tuning of cell weight and cell pseudo-mask kernel size hyperparameters for cell model**
**(A)** Scatter plots of lymphocyte cell counts for predictions vs ground truth are shown for different combinations of point annotation masking kernel size (rows) and cell class weights (columns). The final selected hyperparameters (5x5 kernel, Lymphocyte weight 100, Other cell weight 50, indicated in magenta) had the best overall correlation performance (Pearson R=0.91, CCC=0.90). **(B)** Effect on cell classification F1, precision, and recall of the kernel size and class weight combinations. Although selection of cell weight and pseudo-mask kernel sizes was done against CCC, the selected parameters

also had favorable F1 score characteristics, namely they were most balanced in terms of precision and recall. Each panel shows sublots for two mask kernel sizes, 7 (top) & 5 (bottom). Within each plot the classification F1, precision, and recall are shown across the range of cell weight combinations. The points corresponding to the selected parameters are indicated by the magenta circle, where precision and recall are most balanced.

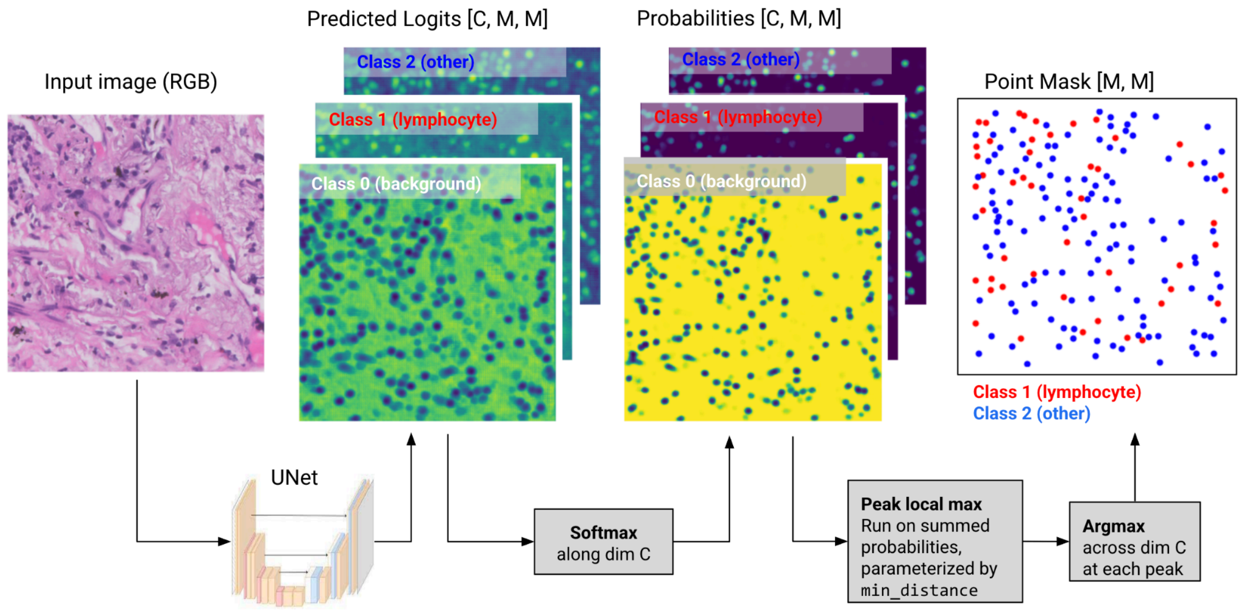


**Figure S6. Method for generating discrete point masks from continuous prediction probabilities**
From left to right: Input RGB images are passed through UNet to produce a 2D logit map per class. Logits are passed through softmax to produce a 2D probability map per class. A peak finding algorithm is used to convert the continuous probabilities to point masks, where each point represents the centroid of a predicted cell. This algorithm is parameterized by a min_distance parameter, which is set to a distance appropriate for lymphocytes.

- Considers a circular neighborhood of (2*min_distance+1)
- For lymphocytes we choose min_distance=5 pixels, which at 20x magnification gives a neighborhood of 6um. Typical lymphocytes are 7-10um in diameter (for example, see https://www.sciencedirect.com/topics/neuroscience/lymphocyte)

Finally, an argmax function is applied to the local peaks across classes (excluding background class) to collapse the into a discrete multi-class point mask.

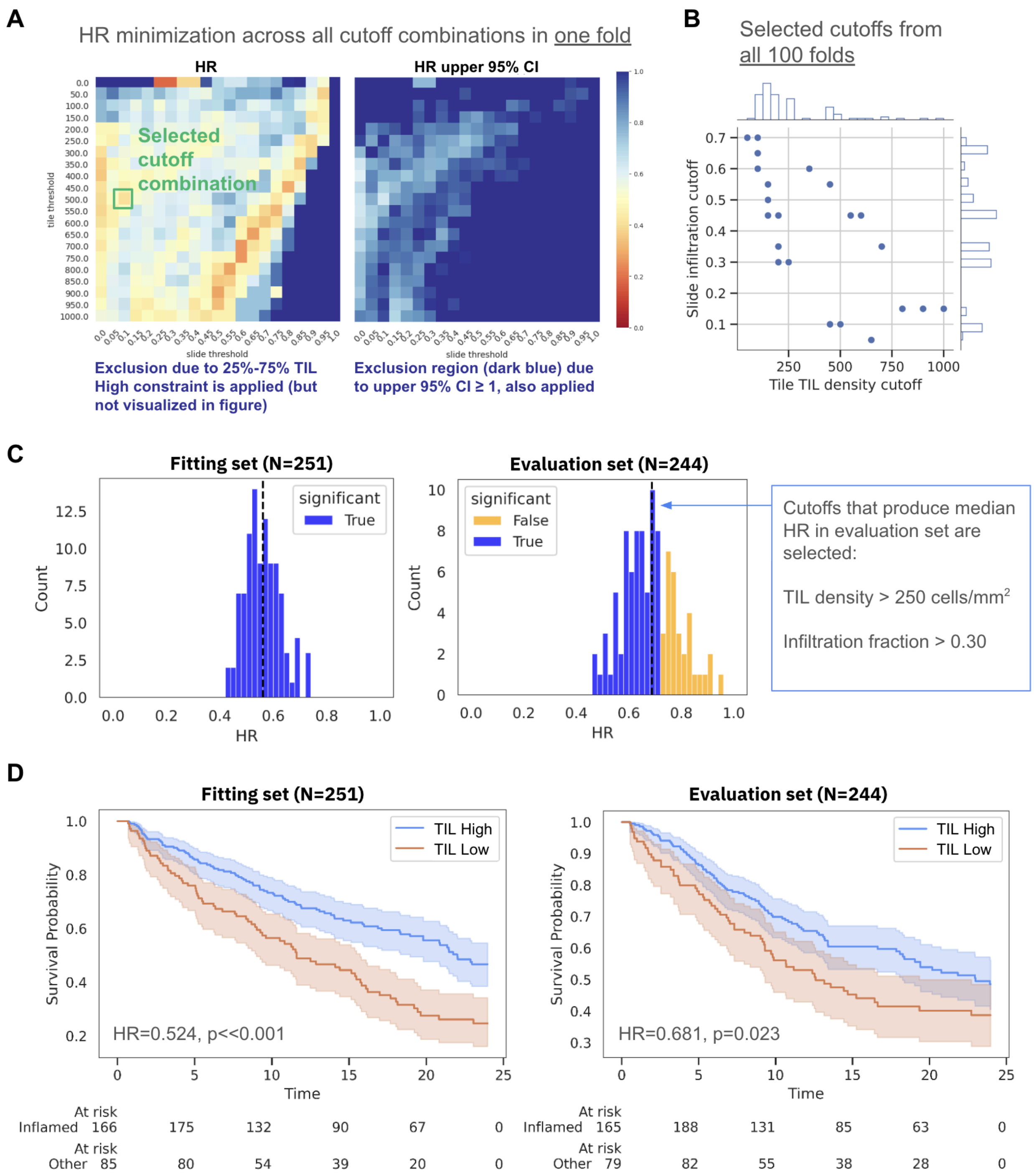


**Figure S7. Cross-validation procedure for selecting tile-level and slide-level cutoffs**

**(A)** Example of constrained HR minimization for one fold. Across 400 combinations of tile-level and slide-level cutoffs, a single cutoff is selected that minimizes the HR (green square) while obeying the constraints. **(B)** From all 100 folds, the selected tile-level (x-axis) and slide-level (y-axis) cutoffs that minimized the HR in the fitting set are shown (each point represents a fold). Marginal distributions are shown where points overlap. Note that the two cutoffs have a commensurate relationship to each other. **(C)** Cross validation HR minimization results are shown for the fitting (left) and evaluation (right) sets. HRs for which the 95% confidence interval exceeded 1 are considered non-significant (orange). While all folds in the fitting set produced significant HR values, roughly 30% of the HR values in the evaluation set were not significant. Tile-level and slide-level cutoffs that produce median HR in the evaluation set are selected, namely tile-level TIL density > 250 cells/mm$^2$ and slide-level infiltration fraction > 0.30. **(D)** For the selected cutoffs, Kapplan-Meier stratification is performed on fitting (left) and evaluation (right) sets, where TIL High is defined as slide-level infiltration fraction > 0.30.

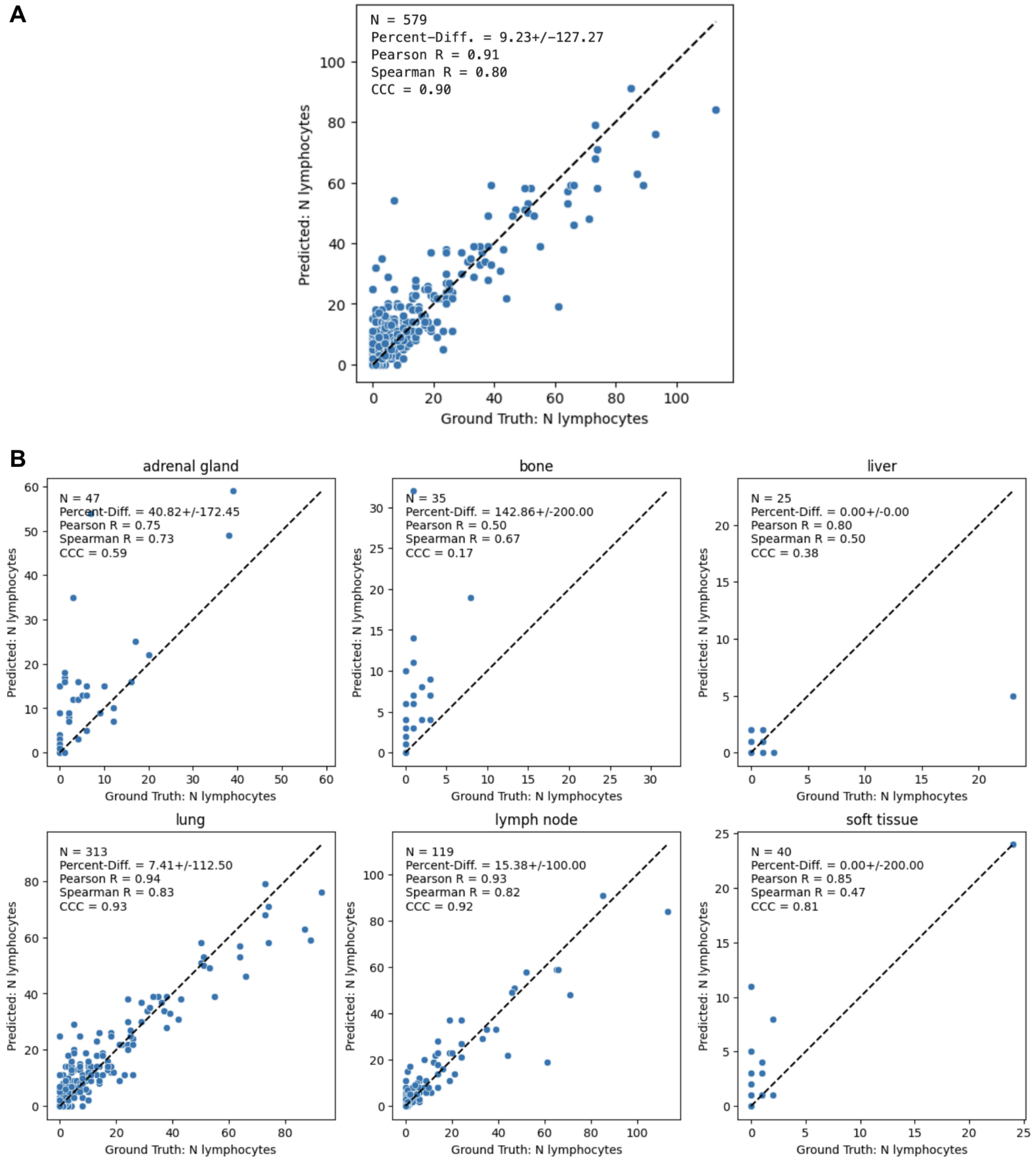


**Figure S8. Scatter plots of per-FOV lymphocyte cell counts in predictions vs ground truth.**

**(A)** Scatter plot of prediction vs ground truth across the entire analytical test set (CCC=0.90), with each point representing the lymphocyte count within an FOV. **(B)** Scatterplots grouped by tissue site. Predictions are close to the x=y unity line for lung (CCC=0.93) and lymph node (CCC=0.92) tissue sites. The model overpredicts relative to ground truth in adrenal gland, bone, and soft tissue. Liver tissue exhibits extremely low cell counts, making the CCC metric unstable for that condition.

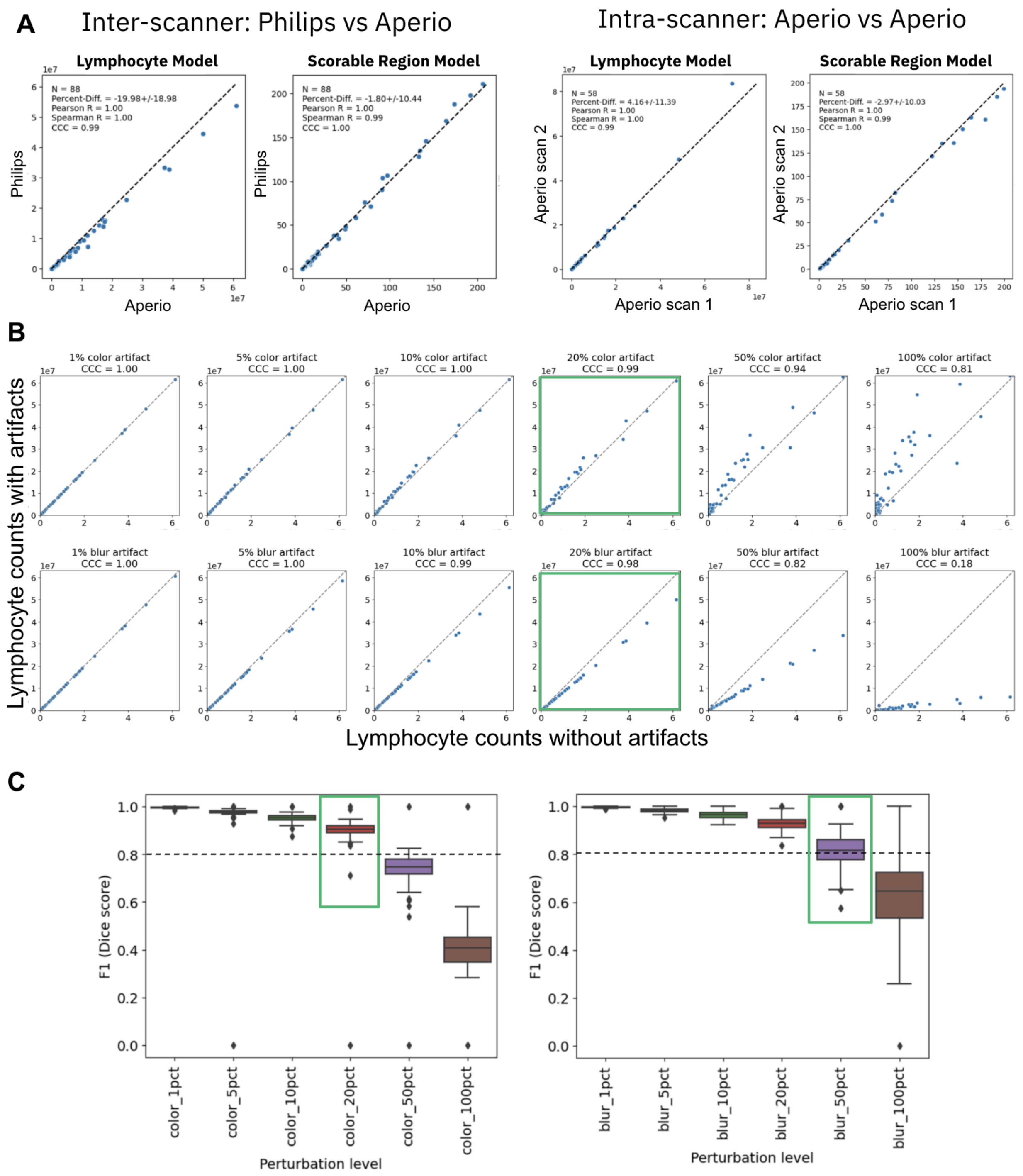


**Figure S9. Model robustness to scanner differences and simulated color and blur artifacts.**
**(A)** Lymphocyte and scorable region model predictions are nearly identical between Leica Aperio GT450 and Philips UFS scanners (left) and between rescans on Leica Aperio GT450 scanners (right). **(B)** Lymphocyte detection stability, assessed by the Concordance Correlation Coefficient (CCC) between predictions on unperturbed and perturbed images. CCC remains >0.95 at 20% perturbation for both

artifacts. **(C)** Scorable region segmentation stability, assessed by Dice score. Agreement remains >0.8 at 20% perturbation for color augmentation and up to 50% for blur artifacts.

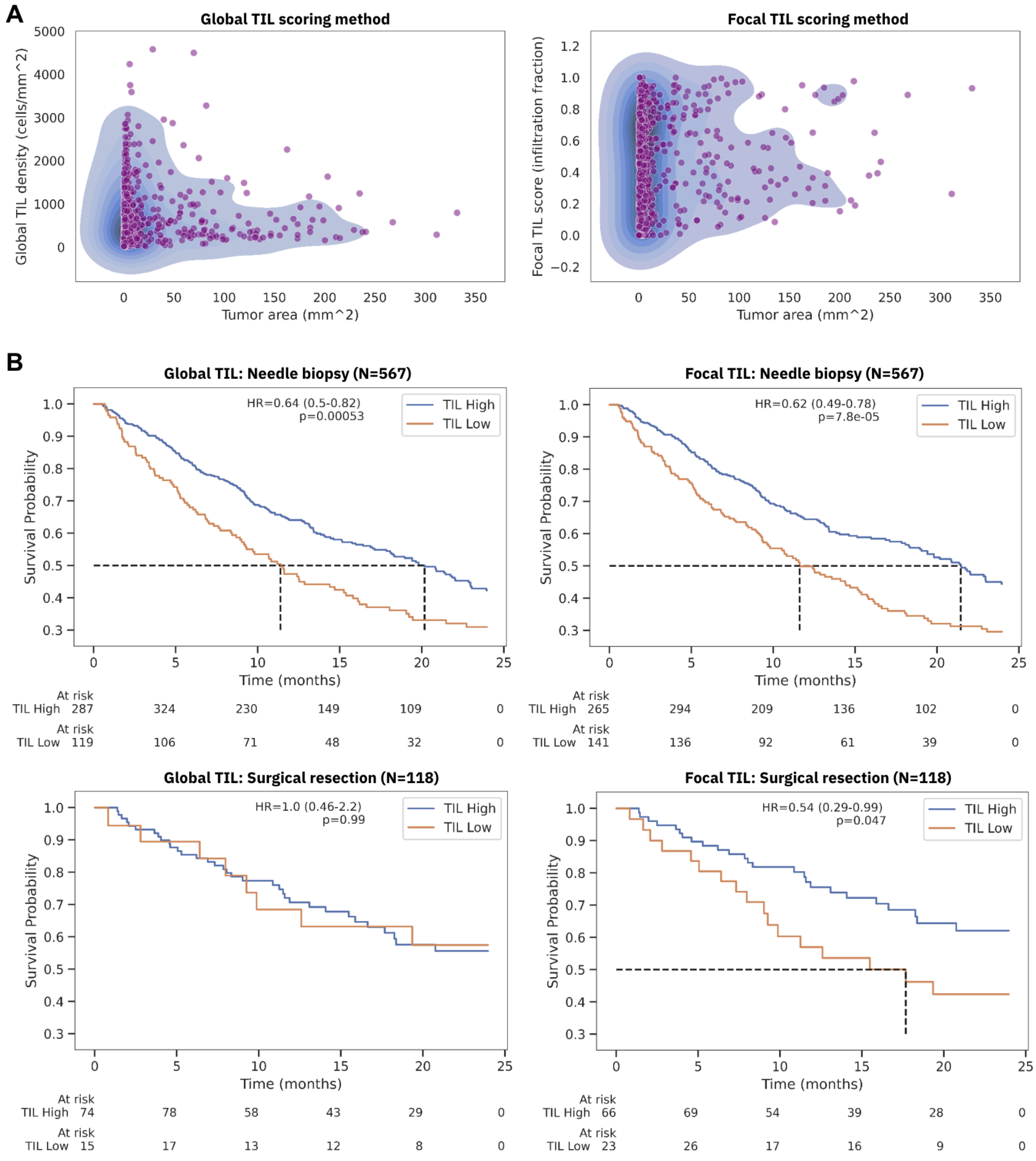


**Figure S10. Advantage of focal TIL score over "global averaging" TIL score**

To demonstrate the advantage of the focal TIL method over other common TIL quantification methods we also implemented a "global" TIL score, where TIL density is measured across the entire tissue as (sum of lymphocytes in scorable region) / (area of scorable region). **(A)** While these methods both capture similar information, the global method tends to under-predict TIL density in larger tissues, such as surgical resections, where lymphocytes cluster in localized regions. In the scatter plots, the distribution of global TIL densities tends to decrease with tumor area, but the distribution of focal TIL score (measured by infiltration fraction) remains relatively constant across tumor areas. **(B)** Using the same TIL density cutoff of 250 cells/mm$^2$ for global and focal TIL methods we measure the prognostic performance of the two methods within procedure type subgroups using Kaplan-Meier analysis. While the focal TIL score significantly stratified OS in both needle biopsies and surgical resections (Right), the global TIL score fails to stratify OS in surgical resections (Left).

### Supplementary Text

##### S.1 TMB definition

Genomic profiling was performed using the Tempus xT DNA sequencing assay. The cohort included samples analyzed using the current xT.v4 panel (648 genes; 1.9 Mb coding region) and the prior xT.v2 panel (596 genes; 2.4 Mb). TMB was calculated as the number of nonsynonymous variations divided by the respective panel size. Included variants were non-silent somatic coding mutations (missense, indel, and stop-loss) with coverage >100x and allelic fraction >5%. To ensure comparability across the cohort, xT.v2 scores were harmonized to the xT.v4 scale. This assay-based TMB calculation is highly correlated with whole-exome sequencing measurements from TCGA ($R=0.986$, $P < 2.2e-16$). For all analyses, TMB-High was defined as an adjusted score of ≥10 mut/Mb.

##### S.2 Algorithm for assigning predicted cells to ground truth cells

First, define an **assignment distance *d*** (i.e. 6 pixels at 20x magnification)

Iterate over each cell in GT. For i'th GT cell:

- Find the nearest predicted cell within ***d***.
- If pred cell is same class as GT → assign on-diagonal **TP**
- If pred cell is different class → assign off-diagonal **misclassification**
    - (i.e. lymphocyte classified as other)
- If no predicted cell within ***d*** → assign **FN**
    - (i.e other classified as background)
- Delete assigned predicted cell from prediction mask

For all remaining predicted cells with no corresponding GT → assign **FP**

- (i.e. background predicted as other)

TN (IGNORED)

- Do not store **TN**, i.e. points where both GT and prediction are background value 0

##### S.3 F1 score from confusion matrix

Following the assignment algorithm detailed in S.2, we define 2 confusion matrices: 1) detection and 2) classification

| **Detection CM** | | Predicted | |
|---|---|---|---|
| | | cell | Background |
| Ground truth | cell | TP | FN |
| | Background | FP | Ignored* |

* Ignored because we do not count TN. Does not contribute to any derived metrics.

| Classification CM | | Predicted | |
|---|---|---|---|
| | | Lymphocyte | Other |
| Ground truth | Lymphocyte | TP | FP or FN* |
| | Other | FP or FN* | TP |

* FP or FN depends on whether computing metric for Lymphocyte or Other

The Detection CM measures how well cells are detected regardless of class, while the Classification CM measures how well cells are classified out of the set that have been detected.

Precision, Recall, and F1 are then computed as

P = TP / (TP+FP+ε)
= cm.diag() / (cm.sum(dim=0) + ε)
R = TP / (TP+FN+ε)
= cm.diag() / (cm.sum(dim=1) + ε)
F1 = 2*P*R / (P + R + ε)

The epsilon (ε) in the denominator is included to prevent division by 0.

##### S.4 Procedure for selection of tile-level and slide-level cutoffs

In the absence of existing consensus on tile-level TIL density and slide-level TIL infiltration cutoffs, we performed a cross validation procedure simulating 100 data splits into fitting and evaluation sets. For each of the 100 folds, data were split 50/50 via randomized sampling stratified by tissue site, procedure type, scanner make, therapy type, and PD-L1 status. Within a fold, Cox PH models were run on the fitting set for 400 unique combinations of tile-level and slide-level cutoffs. The combination that minimized the HR within the fitting set was selected according to the following constraints:

1. TIL density ranged from 50 to 1,000 in steps of 50, infiltration fractions ranged from 0.05 to 1 in steps of 0.05. This resulted in 20^2=400 possible cutoff combinations that could be selected to minimize HR in the fitting set.
2. The upper limit of the HR 95% confidence interval could not exceed 1. This greatly reduces the range of valid cutoffs.
3. The percentage of samples above the two cutoffs had to be within 25-75% of the fitting set population. This excluded cutoffs at the extremes (e.g. TIL density > 50, infiltration fraction > 0.05), that may yield a global minimum of HR, but do not have practical clinical utility due to the extremely skewed High/Low proportions that would result.

An example of this constrained HR minimization grid search on a single fold of the fitting set is depicted in Figure S7A (selected cutoff combination in green box). This is repeated for all 100 folds, creating a distribution of tile-level and slide-level cutoffs, shown in Figure S7B. These cutoffs were then applied to the corresponding evaluation set where HR was measured. The distribution of HR values for fitting and evaluation sets are shown in Figure S7C, where HR values are significant for all 100 folds of the fitting set, but only significant for ~70% of the folds in the evaluation sets. The pair of cutoffs that most often produced the median HR in the evaluation set were then selected as the final cutoffs to stratify populations into TIL High and TIL Low as follows:

**TIL High**

- Tile-level TIL density cutoff > 250 cells/mm$^2$
- Slide-level infiltration fraction > 0.30

**TIL Low**

- Tile-level TIL density cutoff ≤ 250 cells/mm$^2$
- Slide-level infiltration fraction ≤ 0.30

Figure S7D shows Kaplan-Meier analysis with these cutoffs applied in, where the TIL score significantly stratifies OS in the fitting (N=251, HR=0.524, p<0.001) and evaluation sets (N=244, HR=0.681, p=0.023). However, the prognostic performance on the selected evaluation set is not as strongly associated with OS as in the fitting set, suggesting that the selected cutoffs may not always generalize to unseen datasets.
